# COVID-19 Increases the Rate of Incident Hypertension: A Case-Control Cohort Time-to-Event Study

**DOI:** 10.1101/2025.06.09.25329275

**Authors:** Jeremy D. Goldhaber-Fiebert, Shannon C. Phillips, Kimberley D. Lucas, Donna A. Jacobsen, David M. Studdert

**Affiliations:** Department of Health Policy, Stanford Medical School, Stanford University, Stanford, CA, 94306 USA; Center for Health Policy, Freeman Spogli Institute, Stanford University, Stanford. CA, 94306 USA; California Correctional Health Care Services, Elk Grove, CA, 95758 USA; Stanford Law School, Stanford University, Stanford, CA, 94306 USA

## Abstract

**Background:** Of the hundreds of millions of COVID-19 cases globally, most have been non-fatal, though “Long COVID” after acute infection has been documented in many. While many studies have reported post-COVID increases chronic disease incidence including hypertension (HTN), racial/ethnic minority study participants are underrepresented and studies often lacks controls for potential confounds.

**Methods:** We conducted a case-control cohort time-to-event study of 39,746 individuals incarcerated in 31 California state prisons. The main outcome was incident diagnosed HTN among individuals incarcerated continuously since January 1, 2019 with no HTN diagnosis prior to March 1, 2020 (beginning of the unexposed period of observation). The main exposure was a positive COVID-19 test, with the exposure period beginning 31 days afterwards (post-acute period). Covariates included age, gender, race/ethnicity, BMI, and blood pressure at the pandemic’s start, frequency of healthcare contacts prior to the pandemic, and COVID-19 testing frequency prior to testing positive. We excluded individuals who lacked BMI or blood pressure measurements prior to or during the pandemic or were never tested for COVID-19 along with those who had been prescribed blood pressure-altering medications. We estimated multivariate Cox proportional hazard models: 1) exposure variable and covariates; 2) adding interactions between the exposure and each covariate. We assessed whether confounding due to changes in HTN testing post-COVID could explain our results.

**Results:** COVID-19 infection significantly increased the rate of incident HTN (main effects model HRR: 1.44 [95%CI: 1.32-1.57]; including interactions HRR: 2.05 [1.50-2.79]). If all individuals in our study had a COVID-19 infection, the 2-year cumulative risk of hypertension was 7.1% [5.7%-8.5%] versus 5.0% [4.2%-5.9%] if none had been infected. The largest absolute effects of COVID-19 on HTN incidence were in those with higher BMIs, higher pre-pandemic blood pressure levels and older ages. Our findings remained consistent with different definitions of the post-acute period and to confounding due to changes in HTN testing post-COVID.

**Conclusion:** COVID-19 increases the risk of incident hypertension long after acute infection. As so many people have had COVID-19 and elevated blood pressure, additional provider awareness and clinical consideration are warranted.

## Introduction

The vast majority of the hundreds of millions of COVID-19 cases globally have been non-fatal, although many involved symptomatic disease, hospitalization, and ongoing comorbidity (1). Understanding of the long-term health effects of COVID-19 remains incomplete (2). “Long COVID”, a wide range of symptoms and conditions that endure for more than 3 months after SARS-CoV-2 infection, is prevalent among US adults, and approximately one quarter of those who experience it report substantial activity limitations (3-4). Furthermore, many studies and meta-analyses have reported post-COVID increases in the incidence of chronic diseases, including diabetes mellitus (DM), cardiovascular, and neurological conditions (5-7).

The current evidence base for the increase in incidence of hypertension (HTN) post-COVID has several limitations (5-7). First, despite experiencing disproportionately high rates of both COVID-19 and HTN in the US, minorities, particularly Hispanic and non-Hispanic Black individuals, are underrepresented in many of these studies, limiting generalizability. Second, many studies have small sample sizes, reducing the precision of their estimates and limiting their ability to examine specific subpopulations. Third, some studies were not designed to systematically ascertain and exclude baseline, pre-existing HTN, or confounding medications that may elevate blood pressure levels.

Finally, most of the studies were vulnerable to potential confounding – either from lack of systematic and frequent testing for both the exposure (COVID-19) and outcome (HTN), or the absence of adjustment for baseline levels in elevated blood pressure (below diagnostic thresholds) or changes in rates of HTN diagnostic ascertainment after COVID-19.

In this case-control cohort time-to-event study, we used data on 39,746 individuals continuously incarcerated in California’s state prison system before and during the pandemic. Our goal was to examine the extent to which a first infection with SARS-CoV-2 increased the risk of HTN. The large, racially and ethnically diverse nature of this population study extends the prior literature. Further, thanks to CDCR’s policies, internal health system, and electronic health records, the prison residents received frequent and systematic testing for both SARS-CoV-2 and HTN, and there was systematic tracking of both prior diagnoses and potentially confounding medications.

## Methods

### Data

The California Correctional Health Care Services (CCHCS), which delivers healthcare services to California’s incarcerated population in 31 prisons that California Department of Corrections and Rehabilitation (CDCR) oversees and operates, provided data on each resident in these prisons. Data included demographic information (e.g., age, race and ethnicity, and gender) and detailed clinical information in an electronic health record that contains dates and types of clinical encounters, anthropometric measures, results of physical examinations, medication prescriptions and dispensing, and laboratory tests and results. Importantly, the systematic testing program for incarcerated residents during the pandemic period provided information on all sample collection dates and detailed results of all reverse-transcriptase-polymerase-chain-reaction (RT-PCR) and antigen SARS-CoV-2 tests.

CDCR and CCHCS partnered with Stanford University to establish a deidentified data feed with the above elements to study the longer-term effects of COVID-19. Stanford University’s Institutional Review Board (IRB-55835) and CCHCS’s Data Access Committee approved this study.

### Participants, Design, and Key Measures

We identified 71,485 individuals who were incarcerated continuously in CDCR prisons during a 14-month pre-pandemic period (January 1, 2019 through February 29, 2020) and then for at least one day during the pandemic period (beginning March 1, 2020). Of these eligible individuals, we excluded any with one or more of the following characteristics: prior HTN diagnoses, not tested for high blood pressure in the pre-pandemic period, history of confounding medications (see **Appendix S1**), no recorded body mass index (BMI) measurement in the pre-pandemic period, and not tested at least once for both COVID-19 and HTN during the pandemic period. The resultant study cohort comprised 39,746 individuals (**Appendix Figure 1**).

We conducted a case-control cohort time-to-event study. The primary outcome of interest was time to incident HTN, which we coded as occurring on the earliest date a diagnosis, healthcare claim (for those incarcerated individuals seen by a provider outside of the CDCR medical system), or medication for HTN was recorded in the medical record (see **Appendix S2** for further details). A high recorded blood pressure measurement without a recorded diagnosis, claim, or medication was not considered an HTN outcome as it alone would not satisfy the diagnostic criteria for HTN. The exposure of interest is SARS-CoV-2 infection. To focus on time after acute infection, the exposure period of interest began on the 31^st^ day after the first collection date for a sample that tested positive for SARS-CoV-2.

All cohort members began by contributing (non-exposure) observation time. Individuals who had their first positive COVID-19 test prior to being diagnosed with HTN began contributing exposure time at 31 days after this test sample was collected (we excluded the first 30 days from the analysis). The following were censoring events, whether they occurred during exposure or non-exposure time: death, release from prison, positive COVID-19 test (based on collection date), starting a confounding medication, and the end of observation time (March 1, 2023).

### Other Measures

Our analyses adjusted for age, gender, and race and ethnicity, all of which have been linked directly or indirectly to differential risk for HTN in previous studies (8). Residents’ ages on March 1, 2020 were categorized as 18-39 years, 40-59 years, or 60+ years. Residents’ gender was classified as male or female per administrative records.

Residents were classified into one of four racial or ethnic groups: Hispanic, non-Hispanic Black, non-Hispanic White, and Other (see **Appendix S3**).

We adjusted for healthcare system use patterns and baseline clinical characteristics that may be associated with COVID-19 or HTN—either with risk of developing these conditions or with the likelihood and timing of being detected with them. The healthcare use variables included the frequency of days with 1+ clinical contact in the pre-pandemic period (<4 contact-days/year, 4-11 contact-days/year, or 12+ contact-days /year) and the frequency of SARS-CoV-2 testing prior to testing positive (<6 tests/year, 6-11 tests/year, or 12+ tests/year). Baseline clinical characteristics included the most recent pre-pandemic assessment of blood pressure (mmHg) (Normal [systolic <120 and diastolic < 80]; Elevated [systolic 120-129 and diastolic < 80]; High: [systolic ≥130 and diastolic ≥80]) and of body mass index (BMI, kg/m^2^) (Non-Overweight/Obese [<25], Overweight [≥25 and <30], Obese [≥30]) (see **Appendix S3**).

### Statistical Analyses

The primary goal of our analyses was to test for potential changes in the rate of incident HTN caused by a first SARS-CoV-2 infection.

Our main analysis used multivariate Cox proportional-hazards models to estimate this relationship. We first regressed incidence of HTN on positive diagnosis of SARS-CoV-2 infection (i.e., the exposure variable of interest), to calculate hazard rate ratios (HRRs), adjusting for the covariates described above. In a second model, we added interactions of the exposure variable with each covariate. Prior to conducting any analyses, we adopted the following rule to choose between these two models for purposes of secondary analyses and visualizations. We conducted two tests (whether the interaction coefficients are significant; whether the linear combination of the coefficients for the predefined subgroups [COVID main effect coefficient and relevant interaction coefficient] are significant). If neither test showed significance, then select the main-effects-only model, otherwise select the model with interactions.

We computed predicted cumulative hazard curves (with bootstrapped 95% uncertainty intervals) under two scenarios: 1) no one in the CDCR baseline study population was infected with SARS-CoV-2; 2) everyone in the CDCR baseline study population was infected with SARS-CoV-2 (**See Appendix S4**). If the model with interactions was selected based on the rule above, suggesting baseline rates of HTN differed and there was heterogeneity of the effect of SARS-CoV-2 infection on these rates, we predicted these curves for subgroups defined by either age and blood pressure level or by BMI status and blood pressure level, allowing the patterns of other covariates to be as observed for all those included in the analysis from each respective subgroup (**See Appendix S4**).

### Sensitivity Analyses

To probe the robustness of our estimates to our choice of starting the exposure period 31 days after testing positive for SARS-CoV-2 infection, we re-ran our main analyses using three other definitions for the beginning of the exposure period (starting on the same day as the resident first tested positive for SARS-CoV-2 infection; starting on the 61^st^ day after testing positive; starting on the 91^st^ day) (**Appendix Figures 2-4**).

### Assessment of Confounding by Change in Post-COVID HTN Diagnostic Testing Frequency

If the frequency of diagnostic testing for HTN changes after testing positive for SARS-CoV-2, then this can result in a greater apparent rate of diagnosed HTN which would confound the analysis. We performed the following additional analyses to determine whether changes in diagnostic rate after testing positive for SARS-CoV-2 would be sufficient to explain the estimated effect of SARS-CoV-2 on the incidence rate of HTN. First, for individuals who tested positive for SARS-CoV-2, we determined the change in their blood pressure testing rate by computing the difference between the average monthly rate of blood pressure testing for 210-30 days prior to their positive SARS-CoV-2 test and for 30-210 days after their positive SARS-CoV-2 test.

Second, for people who did not test positive for SARS-CoV-2, we computed a “falsification” change in their blood pressure testing rate. Specifically, we estimated quantile regressions (regressions for each quantile from the 1^st^ to the 99^th^) to describe the distribution of the number of days from the start of the pandemic period until the first positive SARS-CoV-2 test for all people in our analysis who tested positive for SARS-CoV-2; our calculations adjusted for baseline age, gender, race/ethnicity, frequency of clinical contact in the pre-pandemic period, body mass index, and frequency of SARS-CoV-2 testing prior to testing positive. These quantile regressions were used to describe inverse cumulative density functions for each combination of the covariates from which we then sampled to impute a “falsification” SARS-CoV-2 positive date for each person with the corresponding pattern of covariates in the study population who did not contribute exposure time. Using this “falsification” date, we computed the change in their blood pressure testing rates pre/post “testing positive for SARS-CoV-2” as described above.

Third, we estimated multivariate linear regressions with change in blood pressure testing rate as the outcome and whether the person had actually tested positive for SARS-CoV-2 as the main predictor, adjusting for the same covariates as in our main analyses. We interpreted a positive coefficient from this regression as the average change in the diagnostic testing rate due to testing positive for SARS-CoV-2. Similarly, we computed the average blood pressure testing rate before/without a positive SARS-CoV-2 test, which enabled us to estimate the proportional change in diagnostic testing rate due to SARS-CoV-2.

Fourth, we repeated the entire procedure for generating the dataset and estimating the effect of testing positive for SARS-CoV-2 on the HTN diagnostic testing rate described up to this point 1,000 times in order to bootstrap uncertainty bounds around the magnitude of our estimate of the change in diagnostic testing rate due to SAR-CoV-2 positivity.

Finally, we used a simulation model that combined estimates from our main analysis of the effect of COVID-19 on HTN incidence and the changes in HTN diagnostic testing due to testing positive for SARS-CoV-2 to compute how large an effect on the apparent rate in HTN incidence could be due to changes in diagnostic testing of the magnitudes we estimated -- and hence whether the estimated effect size from the main analysis was likely be explained solely by such a confound (see **Appendix S5**).

## Results

Of the 39,746 individuals included in the analysis, 21,889 (55.0%) tested positive for COVID-19 prior to being diagnosed with HTN or censored. These individuals contributed a total of 20,886,023 days of pre-exposure observation time, during which 1,550 were diagnosed with HTN, and a total of 11,031,077 days of exposure observation time, during which 981 people were diagnosed with HTN. This corresponded with crude HTN rates of 32.5 per 1,000 person-years among those who experienced COVID-19 infection and 27.1 among the uninfected.

Individuals who tested positive for COVID-19 differed in several observable ways from the overall study population (**Table 1**). On average, they were older (14.5% vs. 12.6% who were ≥60 years) and had lower rates of COVID-19 testing (24.4% vs. 32.0% with ≥12 tests/year).

**Table 1.** Baseline Characteristics of People Contributing Pre-Exposure and Post-Exposure Observation Time.

| Group | Pre/No COVID-19 Exposure |  | Post COVID-19 Exposure |  |
| --- | --- | --- | --- | --- |
| Individuals | 39,746 |  | 21,889 |  |
| HTN Events | 1,550 | (3.9%) | 981 | (4.5%) |
| Age 18-39 | 14,790 | (37.2%) | 7,311 | (33.4%) |
| Age 40-59 | 19,953 | (50.2%) | 11,406 | (52.1%) |
| Age 60+ | 5,003 | (12.6%) | 3,172 | (14.5%) |
| Female | 1,277 | (3.2%) | 735 | (3.4%) |
| Male | 38,469 | (96.8%) | 25,154 | (96.6%) |
| Black | 10,834 | (27.3%) | 5,414 | (24.7%) |
| Hispanic | 19,006 | (47.8%) | 10,713 | (48.9%) |
| White | 7,089 | (17.8%) | 4,120 | (18.8%) |
| Other | 2,817 | (7.1%) | 1,642 | (7.5%) |
| COVID-19 tests, <6/yr | 14,839 | (37.3%) | 9,946 | (45.4%) |
| COVID-19 tests, 6-11/yr | 12,200 | (30.7%) | 6,599 | (30.1%) |
| COVID-19 tests, 12+/yr | 12,707 | (32.0%) | 5,344 | (24.4%) |
| Hist. Medical use, <4/yr | 4,415 | (11.1%) | 2,423 | (11.1%) |
| Hist. Medical use, 4-11/yr | 19,371 | (48.7%) | 10,749 | (49.1%) |
| Hist. Medical use, 12+/yr | 15,960 | (40.2%) | 8,717 | (39.8%) |
| Baseline BMI, Non-Overweight/Obese | 8,982 | (22.6%) | 4,708 | (21.5%) |
| Baseline BMI, Overweight | 19,399 | (48.8%) | 10,853 | (49.6%) |
| Baseline BMI, Obesity | 11,365 | (28.6%) | 6,328 | (28.9%) |
| Normal Baseline BP | 16,728 | (42.1%) | 9,416 | (43.0%) |
| Elevated Baseline BP | 8,324 | (20.9%) | 4,578 | (20.9%) |
| High Baseline BP | 14,694 | (37.0%) | 7,895 | (36.1%) |
| Total Exposure Time (days) | 20,886,023 |  | 11,031,077 |  |
| Mean Exposure Time (days) | 525.5 |  | 504.0 |  |
| SD Exposure Time (days) | 361.7 |  | 279.4 |  |

COVID-19 infection was associated with a significantly higher rate of incident HTN in the survival model that included only the exposure variable and the effects of each covariate (HRR: 1.44 [95%CI: 1.32-1.57]) (**Table 2**). The model showed the incidence rates were significantly higher for older individuals, those with higher pre-pandemic blood pressure levels, those with higher BMIs, and differed by race/ethnicity. It also showed that there were significantly higher rates of incident HTN for those with greater frequency of clinical contacts and higher rates of COVID-19 testing.

**Table 2.**
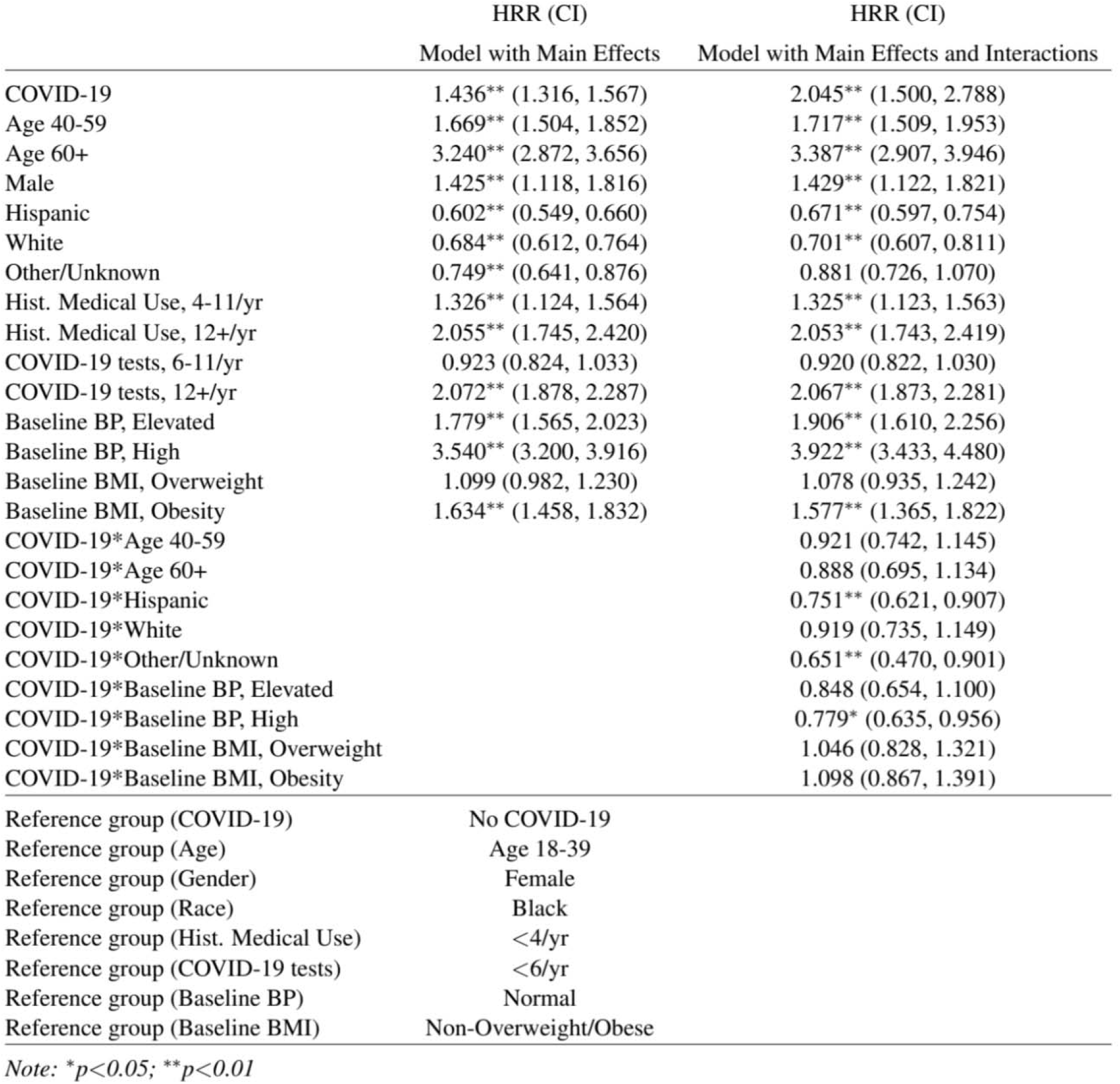
Multivariate Regression Results.

|  | HRR (CI) | HRR (CI) |
| --- | --- | --- |
|  | Model with Main Effects | Model with Main Effects and Interactions |
| COVID-19 | 1.436** (1.316, 1.567) | 2.045** (1.500, 2.788) |
| Age 40-59 | 1.669** (1.504, 1.852) | 1.717** (1.509, 1.953) |
| Age 60+ | 3.240** (2.872, 3.656) | 3.387** (2.907, 3.946) |
| Male | 1.425** (1.118, 1.816) | 1.429** (1.122, 1.821) |
| Hispanic | 0.602** (0.549, 0.660) | 0.671** (0.597, 0.754) |
| White | 0.684** (0.612, 0.764) | 0.701** (0.607, 0.811) |
| Other/Unknown | 0.749** (0.641, 0.876) | 0.881 (0.726, 1.070) |
| Hist. Medical Use, 4-11/yr | 1.326** (1.124, 1.564) | 1.325** (1.123, 1.563) |
| Hist. Medical Use, 12+/yr | 2.055** (1.745, 2.420) | 2.053** (1.743, 2.419) |
| COVID-19 tests, 6-11/yr | 0.923 (0.824, 1.033) | 0.920 (0.822, 1.030) |
| COVID-19 tests, 12+/yr | 2.072** (1.878, 2.287) | 2.067** (1.873, 2.281) |
| Baseline BP, Elevated | 1.779** (1.565, 2.023) | 1.906** (1.610, 2.256) |
| Baseline BP, High | 3.540** (3.200, 3.916) | 3.922** (3.433, 4.480) |
| Baseline BMI, Overweight | 1.099 (0.982, 1.230) | 1.078 (0.935, 1.242) |
| Baseline BMI, Obesity | 1.634** (1.458, 1.832) | 1.577** (1.365, 1.822) |
| COVID-19*Age 40-59 |  | 0.921 (0.742, 1.145) |
| COVID-19*Age 60+ |  | 0.888 (0.695, 1.134) |
| COVID-19*Hispanic |  | 0.751** (0.621, 0.907) |
| COVID-19*White |  | 0.919 (0.735, 1.149) |
| COVID-19*Other/Unknown |  | 0.651** (0.470, 0.901) |
| COVID-19*Baseline BP, Elevated |  | 0.848 (0.654, 1.100) |
| COVID-19*Baseline BP, High |  | 0.779* (0.635, 0.956) |
| COVID-19*Baseline BMI, Overweight |  | 1.046 (0.828, 1.321) |
| COVID-19*Baseline BMI, Obesity |  | 1.098 (0.867, 1.391) |
| Reference group (COVID-19) | No COVID-19 |  |
| Reference group (Age) | Age 18-39 |  |
| Reference group (Gender) | Female |  |
| Reference group (Race) | Black |  |
| Reference group (Hist. Medical Use) | <4/yr |  |
| Reference group (COVID-19 tests) | <6/yr |  |
| Reference group (Baseline BP) | Normal |  |
| Reference group (Baseline BMI) | Non-Overweight/Obese |  |

Note: \* $p < 0.05$ ; \*\* $p < 0.01$

COVID-19 infection was also associated with a significantly higher rate of incident HTN in the alternative survival model that added interactions between the exposure variable and each covariate (HRR: 2.05 [1.50-2.79]). The relationships between each covariate and incident HTN estimated in the model only included main effects were preserved in the model with interactions (**Table 2**). Notably, the interactions between COVID-19 and the covariates were also significant, suggesting heterogenous effects of COVID-19 on rates of incident HTN (**Table 2 and Appendix Table 1**).

For the study population as a whole, COVID-19 infection increased the cumulative hazard of incident HTN at both 1 year (from 2.4% [2.0%-2.8%] to 3.4% [2.7%-4.2%] and 2 years of follow-up (from 5.0% [4.2%-5.9%] to 7.1% [5.7%-8.5%]) (**Figure 1; Appendix Table 2**). COVID-19 had larger absolute effects on the cumulative hazard of incident HTN for subgroups of individuals with higher pre-pandemic blood pressure levels and higher BMIs (**Figure 2**; **Appendix Table 3**) and for subgroups of individuals with higher pre-pandemic blood pressure levels and older ages (**Figure 3; Appendix Table 4**).

**Figure 1.**
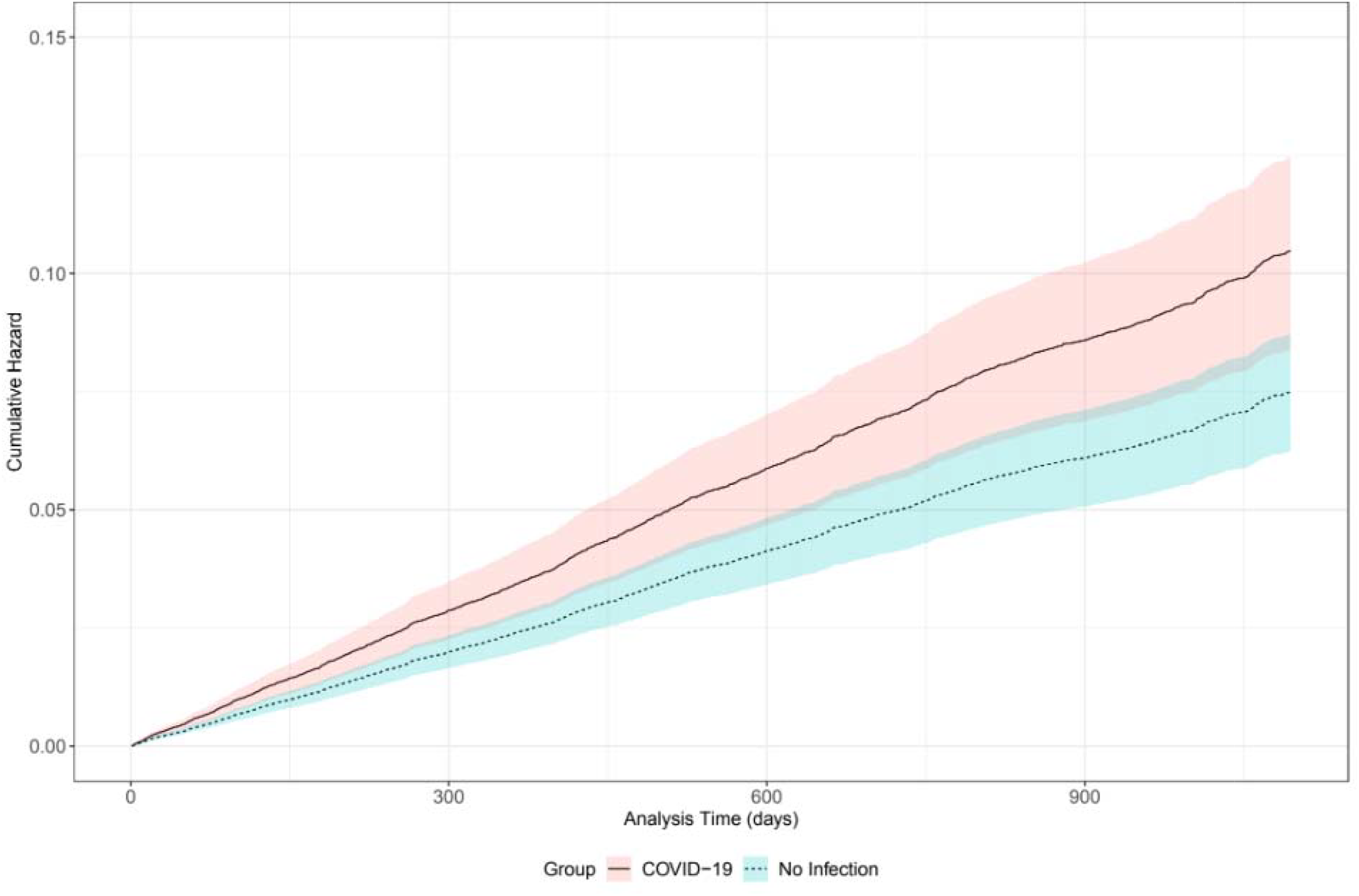
Predicted Cumulative Risks of Incident Hypertension if All Study Participants Had or Had Not Had COVID-19. The Figure shows the predicted cumulative hazard of incident hypertension over time (lines) and associated 95% bootstrapped confidence intervals (shaded regions) of these predictions under two scenarios: a) no one in the study cohort was infected with COVID 19 (dashed line, blue shaded region); b) everyone in the study cohort was infected with COVID-19 (solid line, red shaded region).

**Figure 2.**
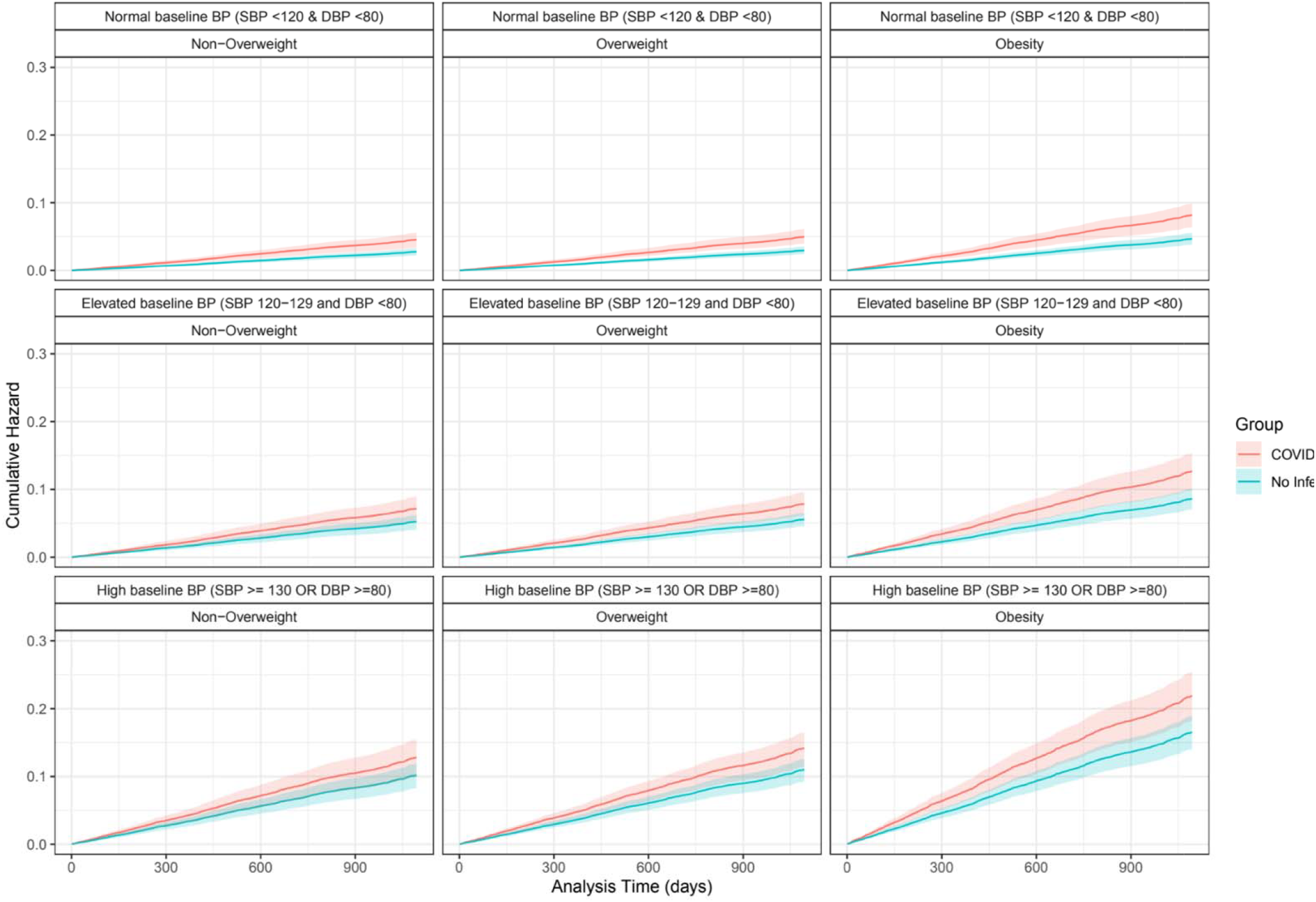
Predicted Cumulative Risks of Incident Hypertension if All Study Participants with Combinations of Baseline Blood Pressure and BMI Categories Had or Had Not Had COVID-19. The Figure shows the predicted cumulative hazard of incident hypertension over time (lines) and associated 95% bootstrapped confidence intervals (shaded regions) of these predictions for subgroups of the study cohort defined in terms of baseline blood pressure and BMI under two scenarios: a) no one in the study cohort subgroup was infected with COVID-19 (blue line, blue shaded region); b) everyone in the study cohort subgroup was infected with COVID-19 (red line, red shaded region).

**Figure 3.**
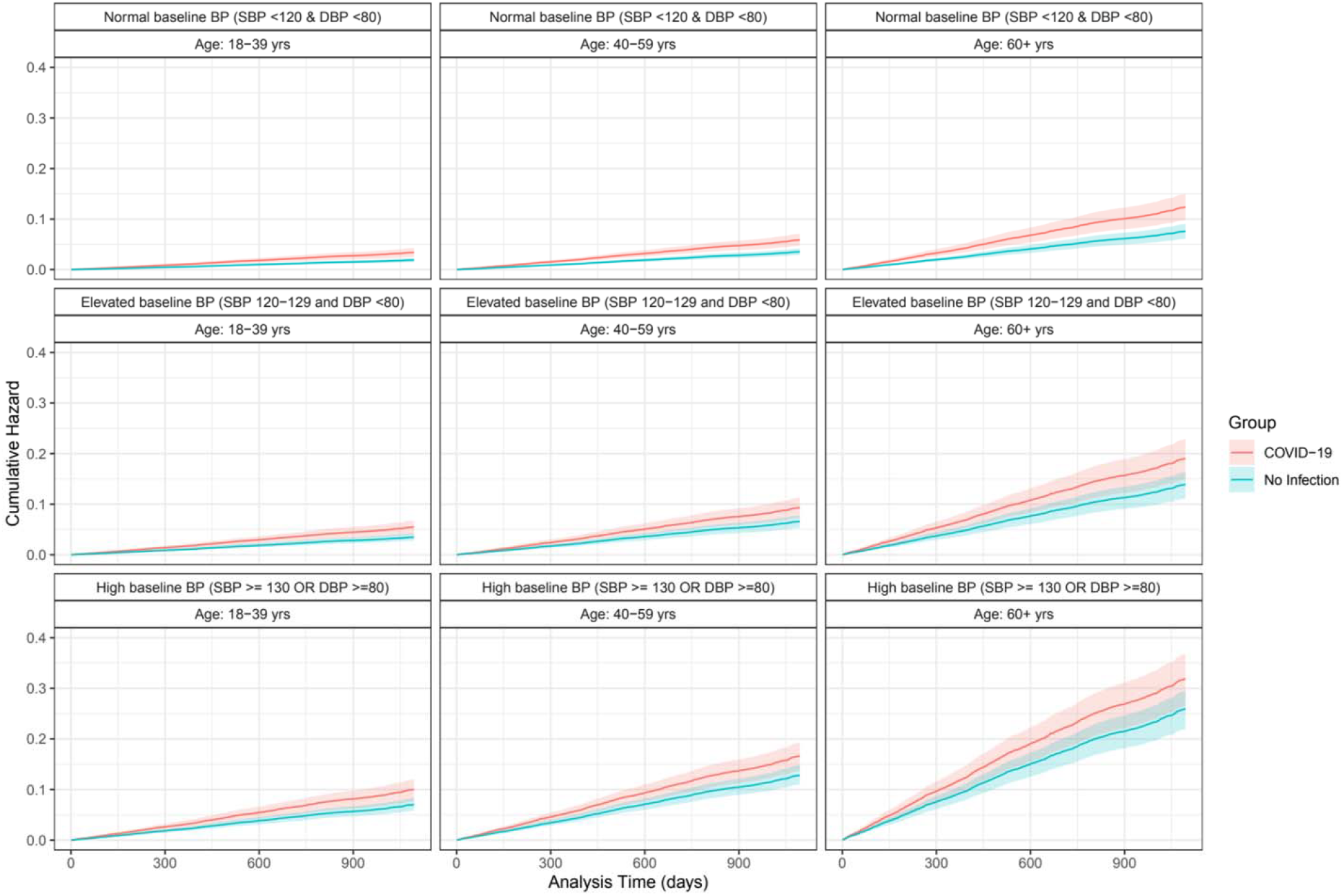
Predicted Cumulative Risks of Incident Hypertension if All Study Participants with Combinations of Baseline Blood Pressure and Age Categories Had or Had Not Had COVID-19. The Figure shows the predicted cumulative hazard of incident hypertension over time (lines) and associated 95% bootstrapped confidence intervals (shaded regions) of these predictions for subgroups of the study cohort defined in terms of baseline blood pressure and age under two scenarios: a) no one in the study cohort subgroup was infected with COVID-19 (blue line, blue shaded region); b) everyone in the study cohort subgroup was infected with COVID-19 (red line, red shaded region).

Our findings remained consistent when we used different cut-offs for the post-exposure observation time (**Appendix Figures 2-4; Appendix Tables 5-8**). Specifically, repeating the main analyses with post-exposure time at 0, 61, and 91 days after the positive COVID-19 test changed the number of individuals analyzed with COVID-19 from 21,889 to 22,293, 21,475, and 21,055 individuals respectively, but the crude rates with COVID-19 remained above 31 per 1,000 person-years, and the direction, magnitude, and significance of the HRRs estimated with both the non-interacted and interacted models were all robust.

In addition, our main findings were robust to tests for potential confounding from variation in HTN diagnostic intensity (**Appendix S5**). Specifically, although rates of clinical contacts in which HTN could have been diagnosed did increase following a positive COVID-19 test by a factor of 1.05-1.25, the magnitude of this increase was not sufficient to explain the estimated effect of COVID-19 on incident HTN in our main analyses. Even taking into account diagnostic confounding in this context, COVID-19 increased incident HTN rates with an estimated HRR adjusted for confounding of nearly 1.40.

## Discussion

Our findings suggest that first infection with COVID-19 increases the rate of developing incident hypertension in a large, racially/ethnically diverse population, especially among individuals aged 40 years and older and those with elevated blood pressure. We also found that the increase in incident hypertension due to COVID-19 persists even after excluding the first 3 months after an acute COVID-19 infection.

These findings contribute in important ways to the existing literature (5-7). Compared with many previous studies of this kind, the population studied -- people incarcerated in the California state prison system -- is larger, more inclusive of younger adults, and more diverse racially and ethnically. The study setting conferred a series of measurement advantages, including rich information on prior diagnoses, healthcare use, and medication use; systematic and frequent COVID-19 testing; and frequent blood pressure testing.

At the same time, generalization from carceral settings, like prisons and jails, to the non-incarcerated population is not straightforward. The levels and prevalence of stressors like confinement, threats of violence, and overcrowding are fundamentally different. Likewise, nutritional differences exist in terms of the quantity and makeup of daily intake. Prior studies have documented the premature aging that incarcerated people undergo (9-10). Hence background rates of hypertension and the magnitude of modification of these rates in older ages may also differ. In addition, the COVID-19 pandemic’s threat to health included infection and mortality rates among incarcerated populations that were 3-5-times higher than in the non-incarcerated population, another unique stressor (11-15). Incarcerated people’s access to exercise and other sources of physical activity were greatly curtailed during periodic episodes of lockdowns (12-13). While both incarcerated people who were and were not infected with COVID-19 experienced these differences, the magnitude of COVID-19’s effects on hypertension could plausibly be more extreme in carceral settings than in non-incarcerated populations. Finally, even though the analysis was conducted in a large carceral system with 10s of prisons housing nearly 100,000 incarcerated people at the start of the pandemic, it should be noted that carceral systems differ in important ways from one another (e.g., shorter terms of incarceration in jails than prisons and often more difficult access to continuity of healthcare) and hence one must be careful in generalizing the magnitude of the estimated effects even to other carceral systems. Despite these caveats, given that people can move between community and carceral settings, sometimes repeatedly, as they are incarcerated and released, our study provides important evidence regarding health risks for 10s of millions of people with current or relatively recent former criminal justice-involvement in the United States.

Our findings have health policy implications and also raise a number of important questions for future research. Our study suggests that systems providing clinical care to this population should be aware that COVID-19 elevates risks of HTN, especially in groups with higher background risks and those in whom COVID-19 appeared to have the largest absolute effects. Likewise, the importance of linkage to health insurance and accessible healthcare upon release is emphasized given the prevalent and elevated risk of HTN in this population (16,17). Additional research is also warranted. It is important to evaluate whether SARS-CoV-2 viral variants all have similar effects on hypertension incidence. Likewise, future studies could evaluate how prior infection as well as vaccination and boosting may modify the increased incidence of hypertension due to COVID-19. It would also be important to determine whether prompt treatment of COVID-19 with anti-viral drugs modifies incident hypertension risk. Finally, studies examining how long after COVID-19 infection elevated incident hypertension risk persists would be particularly important.

In summary, COVID-19 increases the risk of incident hypertension long after the acute infection period. Given the high fraction of people who were infected in both carceral populations and non-incarcerated settings during the pandemic and prevalence of elevated blood pressure, COVID-19’s effect on the population health burden of hypertension could be substantial and warrants increased clinical and population health vigilance.

## Data Availability

All data used in this work were provided by the California Correctional Health Care Services which maintains processes for other researchers to request access.

