## Supplemental Appendices for "COVID-19 Increases the Rate of Incident Hypertension: A Case-Control Cohort Time-to-Event Study"

Shannon C. Phillips, MPH (1,2)

Kimberley D. Lucas, MPH (3)

Donna A. Jacobsen, DO (3)

David M. Studdert, LLB, ScD, MPH (1,2,4)

(1) Department of Health Policy, Stanford Medical School, Stanford University, Stanford, CA, 94306 USA

(2) Center for Health Policy, Freeman Spogli Institute, Stanford University, Stanford. CA, 94306 USA

(3) California Correctional Health Care Services, Elk Grove, CA, 95758 USA

(4) Stanford Law School, Stanford University, Stanford, CA, 94306 USA

**S1. Confounding Medications**

Based on consultations with clinical subject matter experts, medications that can alter blood pressure were identified to enable the determination of clinical events (prescriptions of these medications) that could confound diagnoses of hypertension.

| **GCN Code** | **Description** |
| --- | --- |
| 6758 | betamethasone acetate/betamethasone sodium phosphate |
| 25750, 70506 | budesonide |
| 6780, 6781, 6782, 6784, 6785, 6786, 6787, 6788, 6789, 6790 | dexamethasone |
| 6776, 6778 | dexamethasone sodium phosphate |
| 62053 | dexamethasone sodium phosphate/PF |
| 37045, 6703, 6704, 6705 | hydrocortisone |
| 37045, 6703, 6704, 6705 | hydrocortisone |
| 45311, 6737, 6738, 6741, 6742 | methylprednisolone |
| 6724, 6725 | methylprednisolone acetate |
| 51554, 51555, 51557, 6729, 6730, 6732, 6733 | methylprednisolone sodium succinate |
| 65977, 65978, 65980 | methylprednisolone sodium succinate/PF |
| 45267, 45268, 6746, 6748, 6749, 6750, 6751, 6753, 6754 | prednisone |

**Appendix Figure 1. Flow Diagram of Inclusion in the Case-Control Cohort Time-to-Event Study**

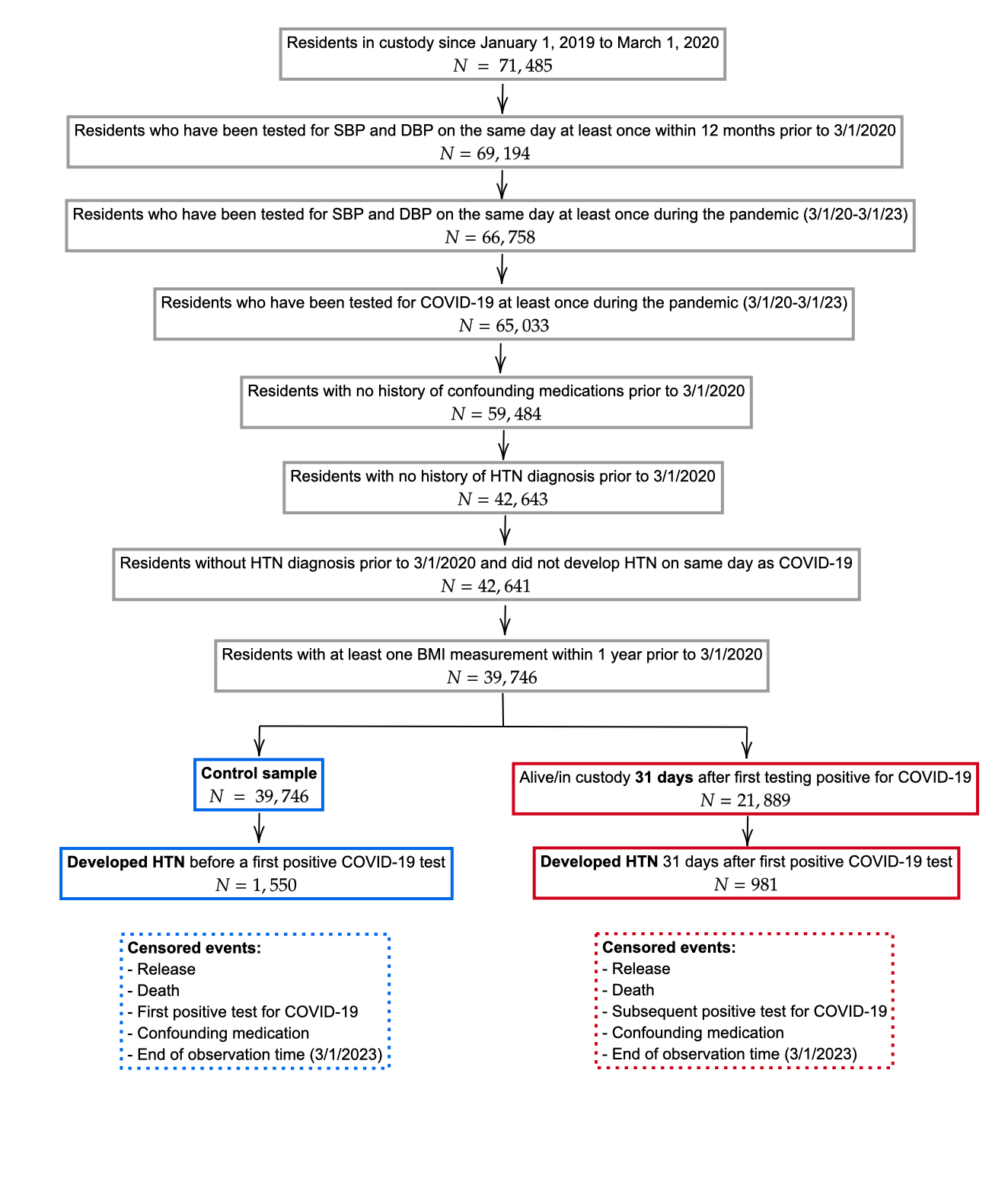

The figure shows the inclusion/exclusion criteria for individual in the study cohort. All cohort members began by contributing (non-exposure) observation time. Individuals who had their first positive COVID-19 test prior to being diagnosed with HTN (a subset of the control/non-exposure sample) began contributing post-exposure time at 31 days after this test sample was collected (we excluded the first 30 days from the analysis).

**Appendix S2. Hypertension Diagnostic Codes**

Individuals were considered to have a hypertension diagnosis if their electronic health record contained any of the following codes. Their diagnosis date was determined as the earliest date at which one of the codes appeared in their health record.

| **Code Type** | **Code** | **Description** |
| --- | --- | --- |
| ICD-10 | H35.03X | Hypertensive retinopathy |
| ICD-10 | I10.xx | Essential (Primary) Hypertension |
| ICD-10 | I11.xx | Hypertensive heart disease |
| ICD-10 | I12.xx | hypertensive chronic kidney disease (CKD) |
| ICD-10 | I13.xx | Hypertensive heart and chronic kidney disease |
| ICD-10 | I15.xx | Secondary hypertension |
| ICD-10 | I16.xx | Hypertensive crisis |
| ICD-10 | I67.4 | Hypertensive encephalopathy |
| ICD-10 | O10.02-O10.03 | Pre-existing hypertension that complicates pregnancy, childbirth, and the puerperium |
| ICD-10 | O13.9 | Gestational [pregnancy-induced] hypertension without significant proteinuria, unspecified trimester |
| IMO | 57871 | Hypertension, essential |
| IMO | 86491 | Hypertension |
| IMO | 193872 | HTN (hypertension) |
| IMO | 1002626 | Mild HTN |
| IMO | 1713042 | Benign essential HTN |
| SNOMed | 193003 | Benign hypertensive renal disease |
| SNOMed | 1201005 | Benign essential hypertension |
| SNOMed | 5148006 | Hypertensive heart disease with congestive heart failure |
| SNOMed | 6962006 | Hypertensive retinopathy |
| SNOMed | 10725009 | Benign hypertension |
| SNOMed | 18050000 | Increased systolic arterial pressure |
| SNOMed | 23154005 | Increased diastolic arterial pressure |
| SNOMed | 24184005 | Elevated blood pressure |
| SNOMed | 28119000 | Renal hypertension |
| SNOMed | 31992008 | Secondary hypertension |
| SNOMed | 32916005 | Nephrosclerosis |
| SNOMed | 38341003 | Hypertensive disorder |
| SNOMed | 38481006 | Hypertensive renal disease |
| SNOMed | 46113002 | Hypertensive heart failure |
| SNOMed | 48146000 | Diastolic hypertension |
| SNOMed | 48194001 | Pregnancy-induced hypertension |
| SNOMed | 50490005 | Hypertensive encephalopathy |
| SNOMed | 56218007 | Systolic hypertension |
| SNOMed | 56265001 | Heart disease |
| SNOMed | 59621000 | Essential hypertension |
| SNOMed | 63287004 | Benign essential hypertension in obstetric context |
| SNOMed | 64715009 | Hypertensive heart disease |
| SNOMed | 65518004 | Labile diastolic hypertension |
| SNOMed | 66052004 | Benign hypertensive heart AND renal disease |
| SNOMed | 66610008 | Malignant hypertensive heart AND renal disease |
| SNOMed | 70272006 | Malignant hypertension |
| SNOMed | 72022006 | Essential hypertension in obstetric context |
| SNOMed | 73410007 | Benign secondary renovascular hypertension |
| SNOMed | 77737007 | Benign hypertensive heart disease with congestive heart failure |
| SNOMed | 78975002 | Malignant essential hypertension |
| SNOMed | 85898001 | Cardiomyopathy |
| SNOMed | 86234004 | Hypertensive heart AND renal disease |
| SNOMed | 89242004 | Malignant secondary hypertension |
| SNOMed | 90493000 | Arteriolar nephrosclerosis |
| SNOMed | 123799005 | Renovascular hypertension |
| SNOMed | 161501007 | H/O: hypertension |
| SNOMed | 170577003 | Good hypertension control |
| SNOMed | 170578008 | Poor hypertension control |
| SNOMed | 194767001 | Benign hypertensive heart disease with congestive cardiac failure |
| SNOMed | 194779001 | Hypertensive heart and renal disease with (congestive) heart failure |
| SNOMed | 194780003 | Hypertensive heart and renal disease with renal failure |
| SNOMed | 194781004 | Hypertensive heart and renal disease with both (congestive) heart failure and renal failure |
| SNOMed | 194785008 | Benign secondary hypertension |
| SNOMed | 194788005 | Hypertension secondary to endocrine disorder |
| SNOMed | 198941007 | Hypertension complicating pregnancy, childbirth and the puerperium |
| SNOMed | 198942000 | Benign essential hypertension complicating pregnancy, childbirth and the puerperium |
| SNOMed | 198944004 | Benign essential hypertension complicating pregnancy, childbirth and the puerperium - delivered |
| SNOMed | 198946002 | Benign essential hypertension complicating pregnancy, childbirth and the puerperium - not delivered |
| SNOMed | 198968007 | Transient hypertension of pregnancy with postnatal complication |
| SNOMed | 233873004 | Hypertrophic cardiomyopathy |
| SNOMed | 307632004 | Non-proteinuric hypertension of pregnancy |
| SNOMed | 371125006 | Labile essential hypertension |
| SNOMed | 371622005 | Elevated blood-pressure reading without diagnosis of hypertension |
| SNOMed | 397748008 | Hypertension with albuminuria |
| SNOMed | 417312002 | Suspected hypertension |
| SNOMed | 428163005 | Hypertensive left ventricular hypertrophy |
| SNOMed | 429198000 | Exertional hypertension |
| SNOMed | 443482000 | Hypertensive urgency |
| SNOMed | 445236007 | Cardiorenal syndrome |
| SNOMed | 697929007 | Intermittent hypertension... |
| SNOMed | 700378005 | Chronic kidney disease stage 3A |
| SNOMed | 700379002 | Chronic kidney disease stage 3B |
| SNOMed | 111411000119103 | End stage renal disease due to hypertension |
| SNOMed | 129151000119102 | Chronic kidney disease stage 4 due to hypertension... |
| SNOMed | 129171000119106 | Chronic kidney disease stage 3 due to hypertension |
| SNOMed | 132721000119104 | Hypertensive emergency |
| SNOMed | 285041000119107 | Benign hypertensive heart disease and chronic renal disease stage 1 |
| SNOMed | 285061000119106 | Benign hypertensive heart disease and chronic renal disease stage 2 |
| SNOMed | 285081000119102 | Benign hypertensive heart disease and chronic renal disease stage 3 |

**Appendix S3. Definitions of Covariates**

**Age:** We defined age at baseline based on the underlying variable BirthYear in the raw data. The difference in years from the start of the pandemic and the birth year produced the age at baseline, which was then grouped as 18-39 years, 40-59 years, 60+ years.

**Sex:** We defined sex as the incarcerated resident’s legal gender as recorded in the raw data at the time of their birth (which is a known limitation for the case of a change of legal gender). This variable does not necessarily correspond to gender identity or morphological sex. Male is coded as 1 and Female as 0.

**Race/Ethnicity:** We defined the race/ethnicity of an incarcerated resident based on the Race variable recorded in the raw data. The raw variable has 10 values (Asian/Pacific Islander (a); Black (b); Cuban (c); D - Pacific Islander (d); Hispanic (h); American Indian/Alaskan Native (i); Mexican (m); Other (o); Unknown (u); White (w)). We map these values to 4 categories:

1. Black (Race = Black)
2. Hispanic (Race in {Hispanic, Cuban, Mexican})
3. White (Race = White)
4. Other/Unknown (Race in {Asian/Pacific Islander, D – Pacific Islander, American Indian/Alaskan Native, Other, Unknown})

**COVID-19 Testing Rate:** We defined the rate of COVID-19 testing prior to a positive COVID-19 as follows. We counted all COVID-19 tests recorded in the raw data with sample collection date prior to the date when the first sample that tested positive was collected and on or after March 1, 2020. For individuals who never tested positive, instead of using the first positive sample date, we used the censor date for this interval. To compute the rate, we divided the number of COVID-19 tests by the number of days in the interval and then multiply by 365 to produce an annual rate.

**Historical Medical Use Rate:** We defined the rate of medical use prior to the start of the pandemic as follows. We counted the total number of medical visits of any type for all days in custody between January 1, 2019 and February 29, 2020 and divided by the number of days in custody in this interval and multiply by 365 to produce an annual rate.

**Baseline BMI:** We wished to categorize body mass index (BMI) of incarcerated individuals as close to the start of the pandemic (March 1, 2020) as possible. To do so, for each incarcerated individual, we obtained maximum height values on all days they are recorded in the raw data. We removed any height values that are outliers (<1.219 meters or >2.134 meters). An individual was then assigned the maximum height value across all his/her/their maximum height values recorded. Then, for each incarcerated individual, we obtained raw maximum weight values (kg) on all days they are recorded in the raw data. We removed any weight values that are outliers (<40.823 kg [90 lbs] or >181.437 kg [400 lbs]). For each day with a weight measurement, we calculated body mass index (BMI) as weight/height^2 (kg/m^2). We then removed outlier BMI values (<16.5 or >50). Finally, for each incarcerated individual, we started from February 29, 2020 and moved backwards day by day (up to 12 months prior) until we identified a day with BMI recorded. We classified this BMI as follows:

1. Non-Overweight/Obese: < 25
2. Overweight: 25 to <30
3. Obesity: 30+

**Baseline Blood Pressure:** We wished to categorize blood glucose of incarcerated individuals as close to the start of the pandemic (March 1, 2020) as possible. For each incarcerated individual, we started from February 29, 2020 and moved backwards day by day (up to 12 months prior) until we identified a day where at least one fasting plasma glucose (FPG) test was recorded. Using this date of the closest FPG test, we then looked back an additional 3 months from this date and identified all FPG tests recorded during this 3-month time window. We then took the average of all FPG tests during this 3-month time window and classified these averages as follows for baseline blood glucose:

1. Normal (SBP < 120 & DBP < 80)
2. Elevated (SBP >= 120 & <130, DBP < 80)
3. High (SBP >= 130, DBP >= 80)

**Appendix S4. Procedures to predict cumulative risks of hypertension for the population and subpopulation included in the analysis**

We predicted the probability of survival (no incidence of hypertension) over time for all combinations of covariates (covariate profiles) from the selected Cox proportional-hazards model using the survfit() function from the “survival” package in R. We then inverted and linearly interpolated the resulting survival curves using the approxfun() function from the “stats” package in R. We generated 5,000 random survival probabilities (uniformly distributed from 0 to 1) to simulate survival times using the interpolated function. This was done for each covariate profile’s survival curve as well as the 95% confidence interval lower and upper bounds. This resulted in each covariate profile having 5,000 simulated survival times (follow up days), as well as the 95% confidence interval lower and upper bounds.

We then computed the weights for each covariate profile by taking the proportion of

unique residents with that covariate profile out of all unique residents in our observed data. There were cases where no residents in our observed data had a particular covariate profile, in which case the weight of an unmatched covariate profile was calculated to be zero.

We then stratified the simulated survival dataset by group status (SARS-CoV-2 Infection vs No Infection). For each stratum, we calculated the proportion of the total weighted population still at risk for incident hypertension out of the total weighted initial population to determine estimated survival probability at each time point (in days) from 0 (where survival probability was set to 1) to the maximum observed follow up time in our data. This was also done for the 95% confidence interval lower and upper bounds for each stratum. To get the final cumulative hazard curve, we plotted 1 - the survival probability estimates (along with the lower and upper bounds) against time in days (from 0 to the maximum observed follow up days) for each stratum (**Figure 1**).

If the Cox model with interactions was selected, we repeated this process of predicting cumulative risks of hypertension by further stratifying each group (SARS-CoV-2 Infection vs. No Infection) by baseline BMI status and baseline blood pressure level (**Figure 2**), as well as age and baseline blood pressure level (**Figure 3**).

For each cumulative hazard curve we predicted, we also extracted the estimated cumulative risk at 1 year (365 days) and 2 years (750 days), along with the 95% confidence interval lower and upper bounds (**Appendix Tables 2-4**).

**Appendix Figure 2. Flow Diagram of Inclusion in the Case-Control Cohort Time-to-Event Study: COVID-19 Test Positive Date Starts Post-Exposure Observation Time**

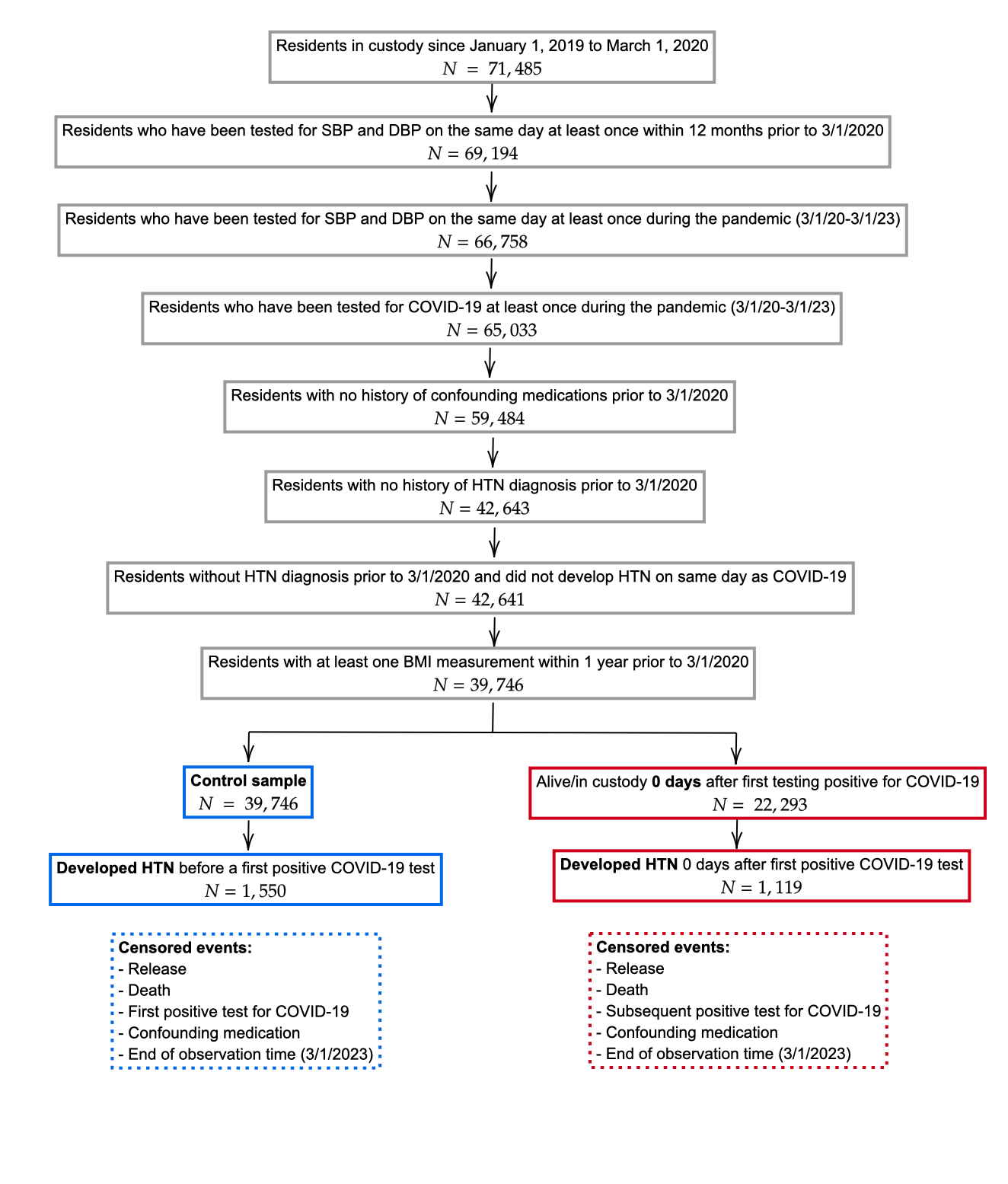

The figure shows the inclusion/exclusion criteria for individual in the study cohort. All cohort members began by contributing (non-exposure) observation time. Individuals who had their first positive COVID-19 test prior to being diagnosed with HTN (a subset of the control/non-exposure sample) began contributing post-exposure time on the day this test sample was collected (we did not exclude any days from the analysis).

**Appendix Figure 3. Flow Diagram of Inclusion in the Case-Control Cohort Time-to-Event Study: 61 Days after COVID-19 Test Positive Date Starts Post-Exposure Observation Time**

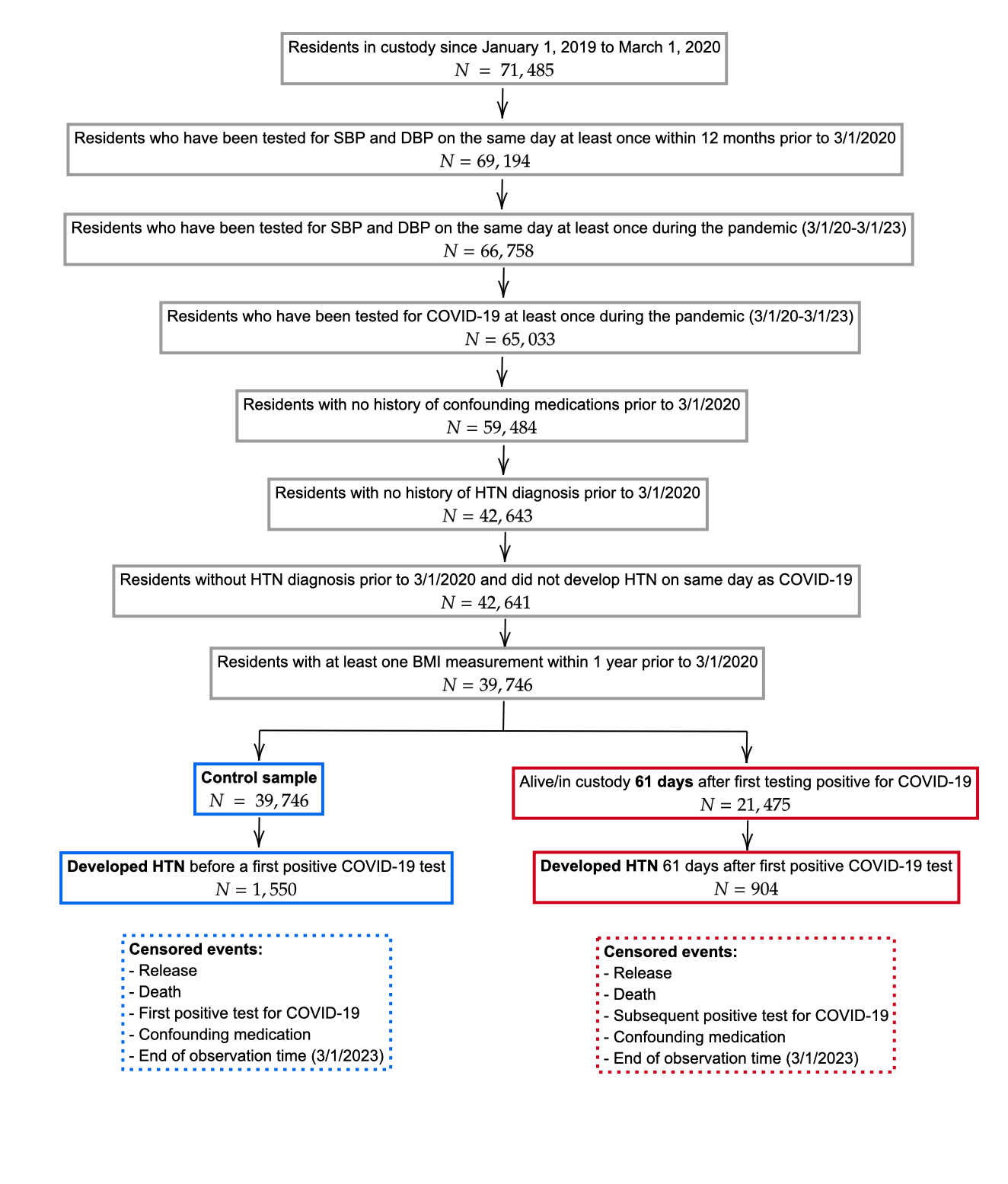

The figure shows the inclusion/exclusion criteria for individual in the study cohort. All cohort members began by contributing (non-exposure) observation time. Individuals who had their first positive COVID-19 test prior to being diagnosed with HTN (a subset of the control/non-exposure sample) began contributing post-exposure time at 61 days after this test sample was collected (we excluded the first 60 days from the analysis).

**Appendix Figure 4. Flow Diagram of Inclusion in the Case-Control Cohort Time-to-Event Study: 91 Days after COVID-19 Test Positive Date Starts Post-Exposure Observation Time**

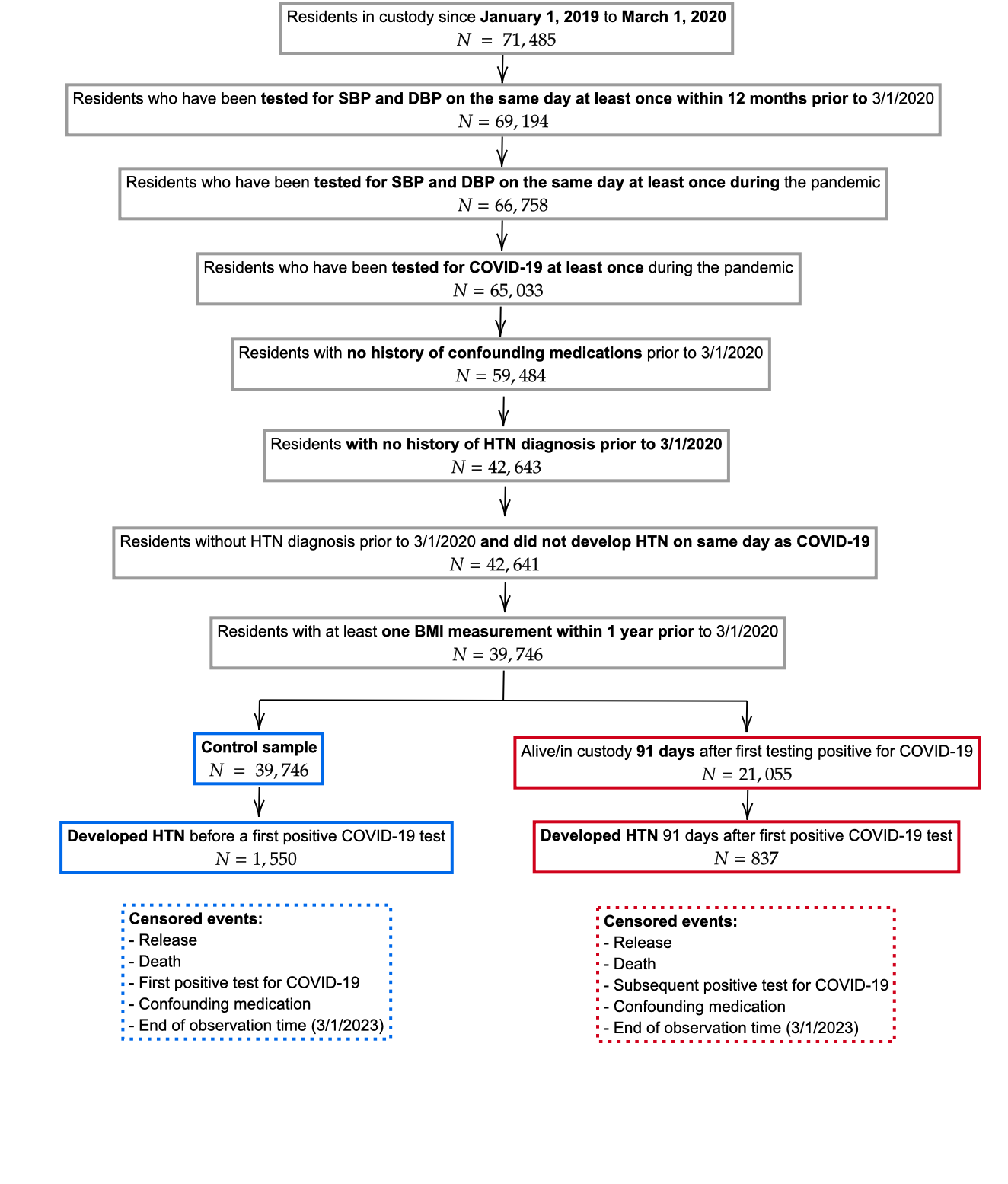

The figure shows the inclusion/exclusion criteria for individual in the study cohort. All cohort members began by contributing (non-exposure) observation time. Individuals who had their first positive COVID-19 test prior to being diagnosed with HTN (a subset of the control/non-exposure sample) began contributing post-exposure time at 91 days after this test sample was collected (we excluded the first 90 days from the analysis).

**Appendix S5. Assessment of the Potential Confounding Effect of Changes in Hypertension Diagnostic Testing after a Positive COVID-19 Test**

Because the outcome we can observe and analyze is incident *diagnosed* hypertension (Dx HTN) and not incident hypertension, it is possible that if COVID infections cause increased rates of HTN diagnostic testing, then observed effects of COVID on Dx HTN incidence could occur even when COVID has no effect on HTN incidence itself. The figure below illustrates this point; Panel A shows a causal model where COVID increases incidence with no effect on diagnostic testing for HTN, and Panel B shows a model where COVID has no effect on HTN incidence but increases rates of diagnostic testing for HTN. We describe the details of our approach for assessing these two possibilities within the context of our analysis below.

*Panel A. An effect of COVID-19 on HTN incidence but no effect on HTN diagnostic testing*

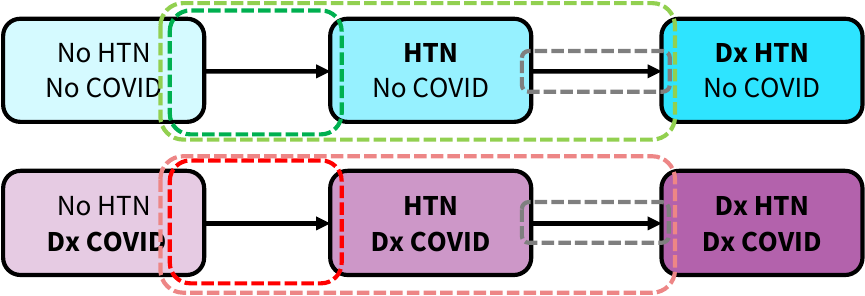

*Panel B. No effect of COVID-19 on HTN incidence but an effect on HTN diagnostic testing*

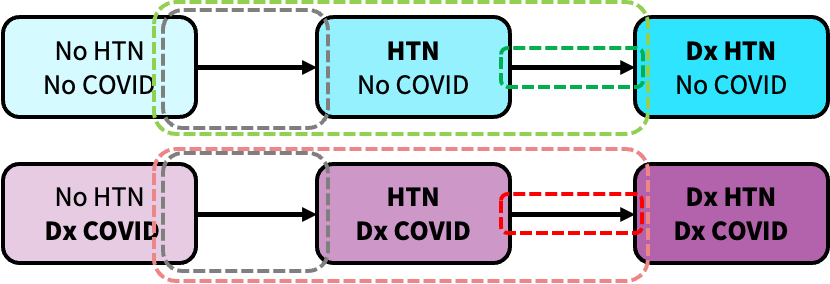

Consistent with the process depicted in the figure, we construct a simple simulation model that includes this process for each of two strata (individuals who have not been infected with COVID and individuals after they have been infected with COVID). In each stratum, there are two rates: 1) the rate of developing HTN ($r_{1}$); 2) the rate of diagnosis of HTN ($r_{2}$). For the stratum for individuals who have been infected by COVID, the model determines these rates with two parameters, thereby relating to the two strata to each other: 1) the hazard rate ratio of HTN for people who have been infected with COVID; 2) the hazard rate ratio of diagnosis of HTN for people who have been infected with COVID.

For the stratum of people who have not been infected with COVID, the cumulative risk of detected hypertension at time *t* is

$$F_{NoInf}\left( t \right)=1-\frac{r_{2}e^{-r_{1}t}-r_{1}e^{-r_{2}t}}{\left( r_{2}-r_{1} \right)}$$

For the stratum of people after they have been infected with COVID, the cumulative risk of detected hypertension at time *t* is

$$F_{COVID19}\left( t \right)=1-\frac{{HRR}_{2}r_{2}e^{-{HRR}_{1}r_{1}t}-{HRR}_{1}r_{1}e^{-{HRR}_{1}r_{2}t}}{\left( {HRR}_{2}r_{2}-{{HRR}_{1}r}_{1} \right)}$$

We apply the simulation model to assess the possibility that confounding due to changes in HTN diagnostic testing rates after a COVID-19 infection could explain the differences we estimated in our main analysis. For example, the main analysis produced estimates of the average 2-year cumulative risk of diagnosed hypertension with and without COVID-19 infection (Appendix Table 2): 5.02% without COVID-19 infection and 7.06% with COVID-19 infection.

In order to do so, we first need to make an estimate of the base rate of HTN diagnostic testing among people who had not tested positive for COVID-19 ($r_{2}$) as well as the hazard rate ratio of HTN diagnostic testing due to testing positive for COVID-19 (${HRR}_{2}$).

Before differentiating between people with or without COVID-19 infections, we observe that the rate of diagnostic testing for HTN (i.e., measuring and recording blood pleasure) varied across people in our sample in the pre-pandemic period and during the pandemic (see table below). On average people were tested slightly more than monthly in both periods, with an increase in the rate of about 2 tests per year during the pandemic period. The 25^th^ and 75^th^ percentiles of testing in both periods were approximately every 2 months to approximately every 3 weeks, again with an increase of about 2 tests per year in both rates during the pandemic.

| Analytic Sample (N=39,746) | 1st Qu. | Median | Mean | 3rd Qu. |
| --- | --- | --- | --- | --- |
| BP test annual rate pre-COVID19 (1/1/2019-2/29/2020) | 5.15 | 9.45 | 13.25 | 15.46 |
| BP test annual rate post-COVID19 (3/1/2020-2/28/2023) | 6.67 | 11.00 | 15.84 | 17.01 |

To estimate the effect of COVID-19 on HTN diagnostic testing rates, we use a differences-in-difference approach. For all people who did not test positive for COVID-19, we assign them a “COVID-19 positive” date. We do this by estimating quantile regressions (1,2,…,98,99^th^ quantiles) of the number of days from the start of the pandemic until the date on which the first positive COVID-19 sample was taken, adjusting for all of the covariates used in our main analysis. The quantiles form estimates of the inverse cumulative density function – if we sample uniformly from 0-1, we can determine between which two quantiles we fall for every individual. Then based on their covariate values we can linearly interpolate these quantiles to determine the day on which they were “COVID-19 positive”. The results of this procedure are shown in the figure below.

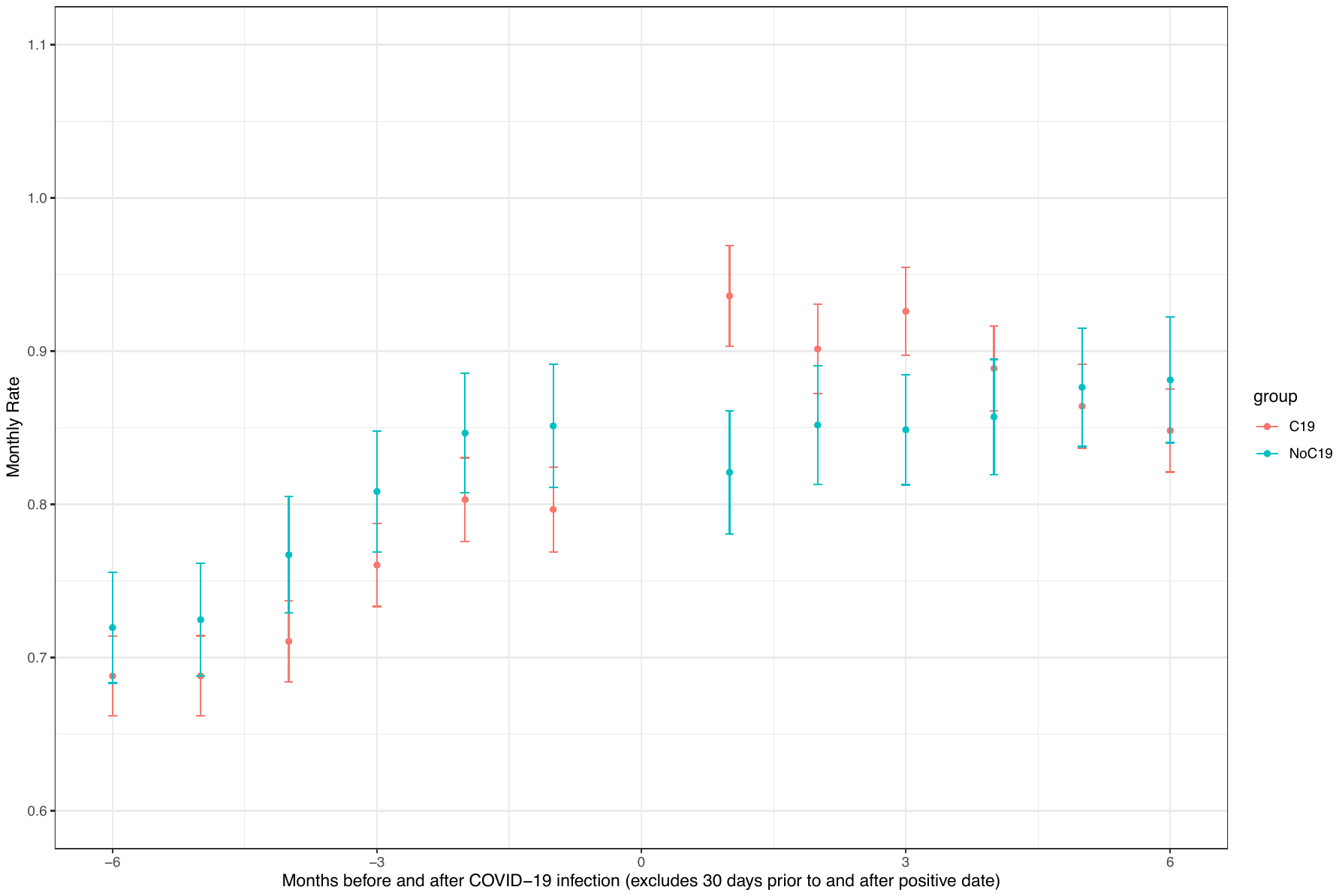

The figure above shows monthly rate 210 to 31 days prior to COVID-19 infection and 31 days to 210 after COVID-19 infection for people who actually had a COVID-19 infection and for those whom we assigned a “COVID-19 positive” date. The crude estimate of the monthly testing rate in the pre-period is 0.83 tests per month in the control group (0.79 in the group of people who had COVID-19 before getting COVID). The crude difference-difference estimate is an increase of about 0.08 tests per month.

|  | Pre | Post | Diff |
| --- | --- | --- | --- |
| COVID-19 | 0.79 | 1.00 | 0.21 |
| Control | 0.83 | 0.96 | 0.13 |
| Diff | -0.04 | 0.04 | 0.08 |

We estimate the difference-in-difference estimate adjusting for baseline covariates and repeat the procedure 1,000 times (i.e., assigning “COVID-19 positive” dates for each person based on random draws based on the quantile regression and then estimating the difference-in-difference regression), producing the results shown in the table below.

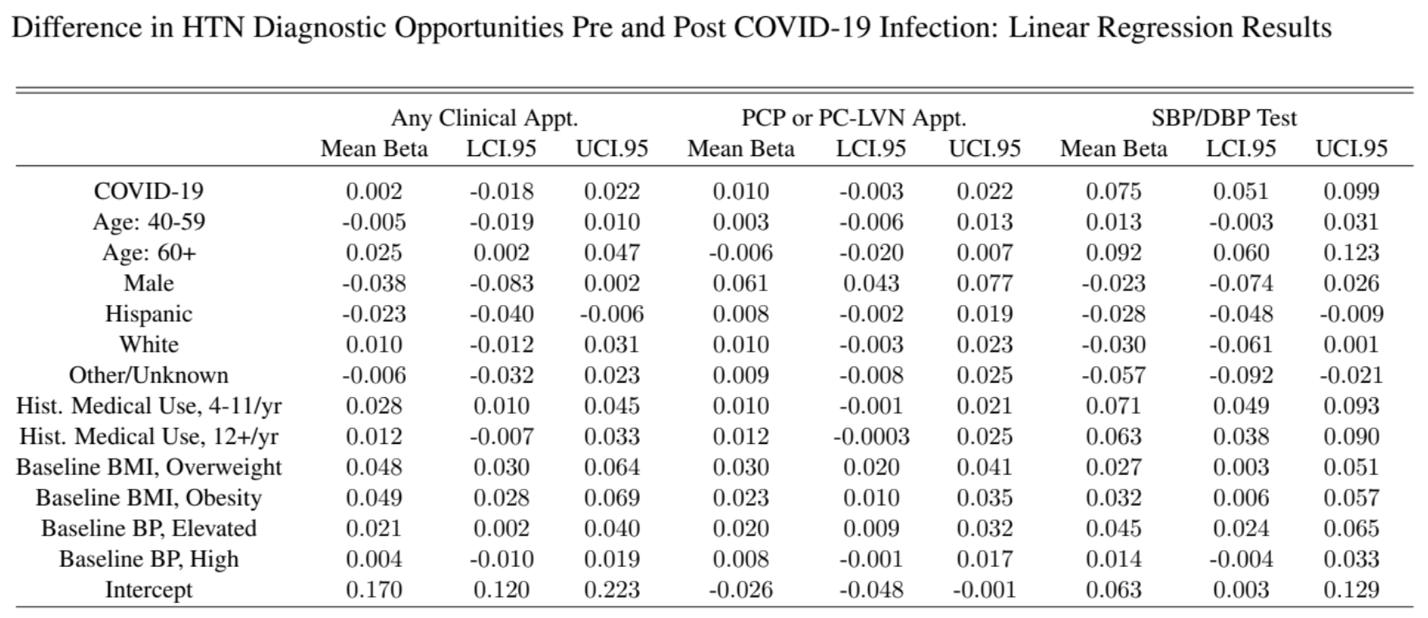

The table above shows results of this procedure for blood pressure testing (SBP/DBP test) as well as for clinical appointments overall and for clinical appointments with healthcare providers who would be most likely to measure blood pressure for the purpose of diagnosis. The results show that a COVID-19 infection did not significantly increase the rate of clinical appointments overall or with particular types of providers (after 30 days post-COVID infection – the post-acute period), but COVID-19 did increase HTN diagnostic testing rates within these visits by 0.075 tests per month [95%CI: 0.051-0.099]. Given the background testing rates of 0.79 to 0.83 in the pre-period, this implies that ${HRR}_{2}$ could be between 1.06 ([0.83+0.051]/0.83) and 1.13 ([0.79+0.099]/0.79) or perhaps at the extreme if all of the change in diagnostic HTN rates were as observed for the COVID-19 group were due to COVID-19 infection itself then it could be 1.27 (1.00/0.79).

The above findings along with our main analyses imply that:

1. $r_{2}$ is likely to be 0.02633 HTN diagnostic tests per day (0.79 HTN diagnostic tests/month)
2. ${HRR}_{2}$ is likely to be about 1.10 and unlikely to be below 1.00 or above 1.27, though we explore values up to 1.50 and even 2.00 in the analyses below.
3. ${HRR}_{1}$ is estimated as 1.44 [1.32-1.57] in our main analysis, and we explore values as low as 1.00 (no effect of COVID-19 on HTN incidence) and as high as 2.00 in the analyses below.
4. $r_{1}$(the rate of incident HTN among those who have not had COVID-19) is not directly observed but cannot be below the rate of incident diagnosed HTN under the assumption that diagnosis is immediate based on the 1-year cumulative rate of diagnosed hypertension in people without COVID-19: 0.000067/day. A reasonable upper bound might be the rate of incident diagnosed HTN among people after COVID-19: 0.000101. In the analyses below we explore, values as low as 0.000060 and as high as 0.00012.

We fixed $r_{2}$ at its observed value and constructed a dense regular 3-dimensional grid of values in the ranges for $r_{1}$, ${HRR}_{1}$, and ${HRR}_{2}$ listed above (1,311,927 unique combinations). We then used $F_{NoInf}\left( t \right)$and $F_{COVID19}\left( t \right)$to simulate the cumulative risks of incident diagnosed HTN at 1 year and 2 years for each of parameter combinations. We computed sum of squared errors of model-simulated values (cumulative risks of incident detected HTN with and without COVID-19 at 1 year and 2 years and the 1-year and 2-year hazard rate ratios if these cumulative risks were converted to rates under an exponential assumption). We computed the inverse of the sum of squared errors for each parameter combination and normalizing these values such that they summed to 1 across the 1,311,927 combinations. We used these values as weights to sample from these combinations with replacement 2,000,000 times.

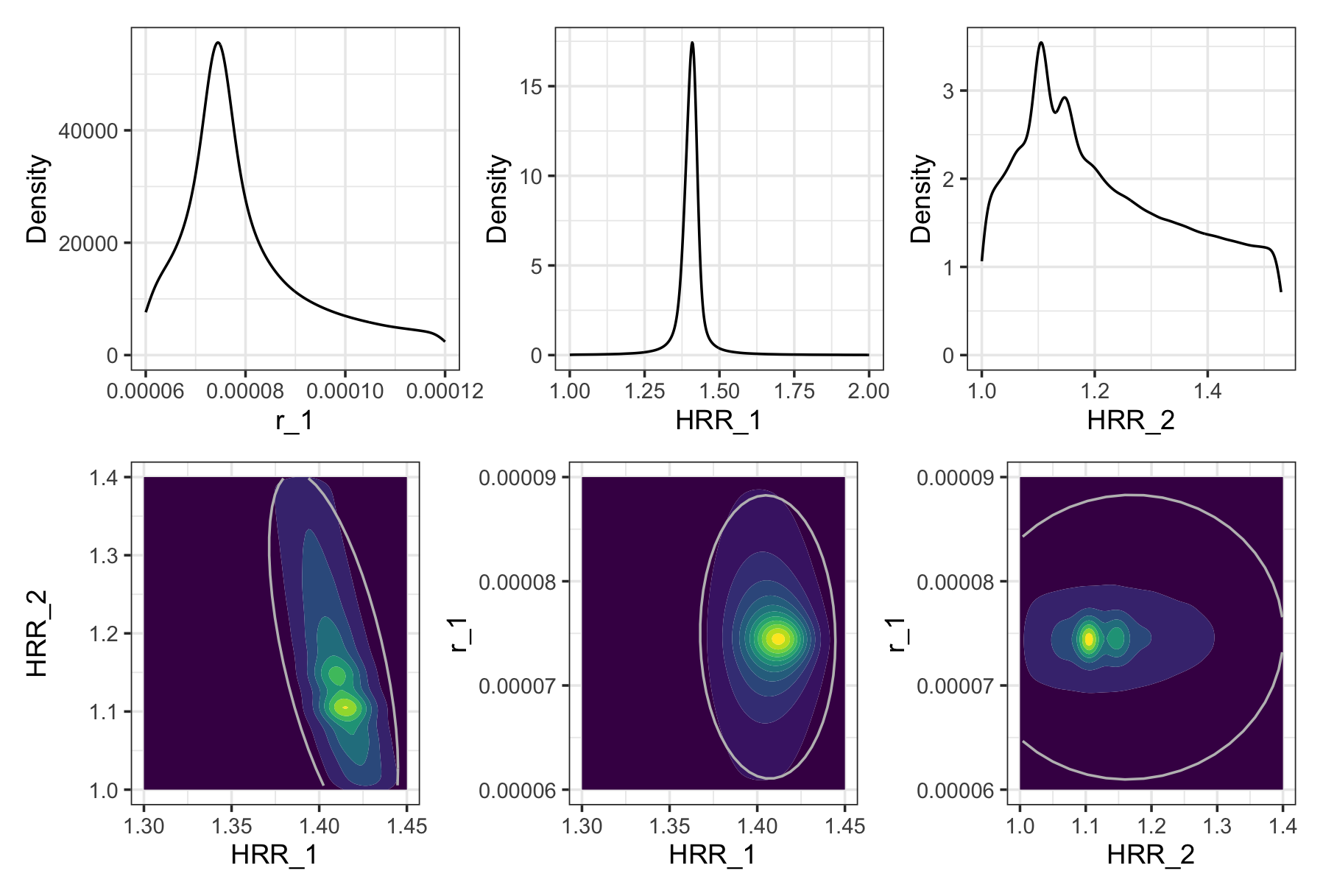

The top row of the figure shows the marginal densities from our procedure – values of $r_{1}$ near 0.000075, values of ${HRR}_{1}$ near 1.40, and values of ${HRR}_{2}$ near 1.12 produce cumulative risks that are most consistent with the observed data. The bottom row shows the relationship between good fitting parameters. In general, if COVID-19 causes a larger increase in the rate of HTN incidence (higher values of ${HRR}_{1}$) it must cause smaller increases in the rate of HTN diagnostic testing (smaller values of ${HRR}_{2}$). Good fitting values of $r_{1}$ are not strongly correlated with values of either ${HRR}_{1}$ or ${HRR}_{2}$. Again, we can see that it is unlikely that the true rate of HTN incidence without COVID-19 is below 0.000065 or above 0.000085 and that is unlikely that the true effect of COVID-19 on the rate of incident HTN is below 1.35 or above 1.45 and that these are consistent with the effect of COVID-19 on the HTN diagnostic testing rate of 1.05 to 1.25.

*First bound: Smallest HTN diagnostic confounding with no effect on incident HTN*

If this entire estimated difference in diagnosed hypertension at is due to differences in HTN diagnostic testing rates, then the rate of incident hypertension ($r_{1}$) for both groups is that observed in the empirical data for those without COVID-19 infection with ${HRR}_{1}=1$ as there is no difference in the rate of HTN incidence between the two groups. Note that we still need to establish $r_{1}$ since we only observe the HTN diagnostic testing rate ($r_{2}$), but this can be easily estimated from our empirical observations and the equation for $F_{NoInf}\left( t \right)$. Finally, we can solve for the smallest value of ${HRR}_{2}$ required to generate the differences in the cumulative risk of diagnosed HTN at 2 years based on $F_{COVID19}\left( t \right)$.

We can then compare the smallest value of ${HRR}_{2}$ required if there is no effect of COVID-19 on HTN incidence to the effect of COVID-19 on HTN diagnostic testing rates we will estimate, and if the smallest value is substantially larger than the estimated effect of COVID-19, we can reject the idea that confounding by HTN diagnostic intensity is sufficient to explain our main results.

If we make ${HRR}_{2}$ twice the upper bound of our estimated effect of COVID-19 on the HTN diagnostic testing rate (>1.50), we find that best fitting values of $r_{1}$ remain centered on 0.000075 and are generally between 0.000065 and 0.000085 and that ${HRR}_{1}$ is very unlikely to be below 1.30 let alone near 1.0. The best fitting combination of values produces model outputs that are consistent with the target data of cumulative risks (shown in the figure below) and have ${HRR}_{1}$ at 1.385.

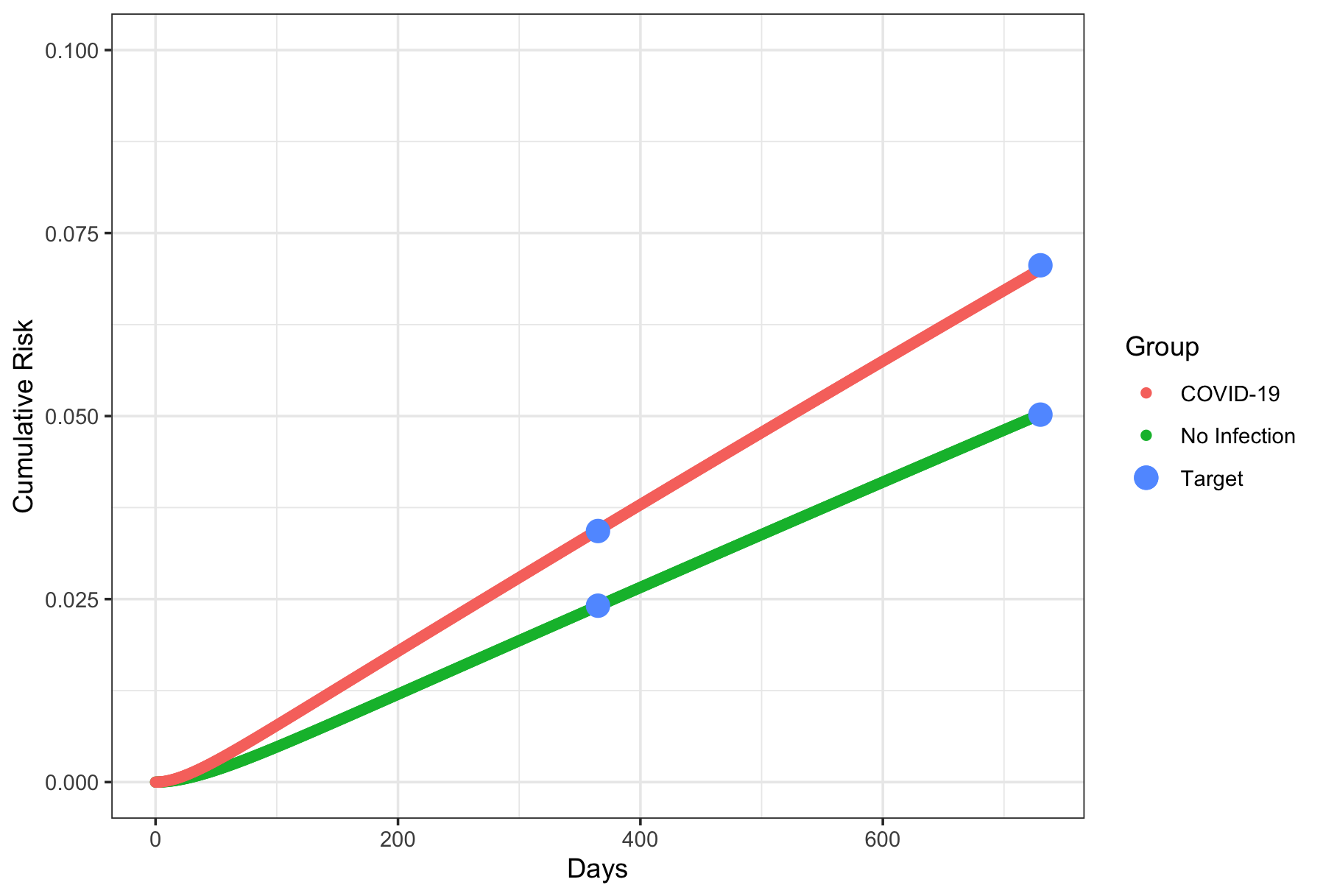

In contrast, when we fix ${HRR}_{1}$ at 1.0 and $r_{2}$ at its observed rate and $r_{1}$ at the value implied for people without COVID-19 (see above) and then search for ${HRR}_{2}$ values as high as 10,000 that cause us to be consistent with the target data of cumulative risks, the best fit we can achieve is shown in the figure below. The reason for this is that HTN diagnostic testing rates are already very high for people without COVID-19 (~ every month to 6 weeks) and thus even if we increase this rate to nearly instantaneous the highest we can get is implied by the underlying rate of incident HTN in people without COVID-19 ($r_{1}$) which is inconsistent with cumulative risks of diagnosed HTN as high as those observed in people after COVID-19. Hence, we conclude that COVID-19 having no effect on underlying HTN incidence is extremely unlikely.

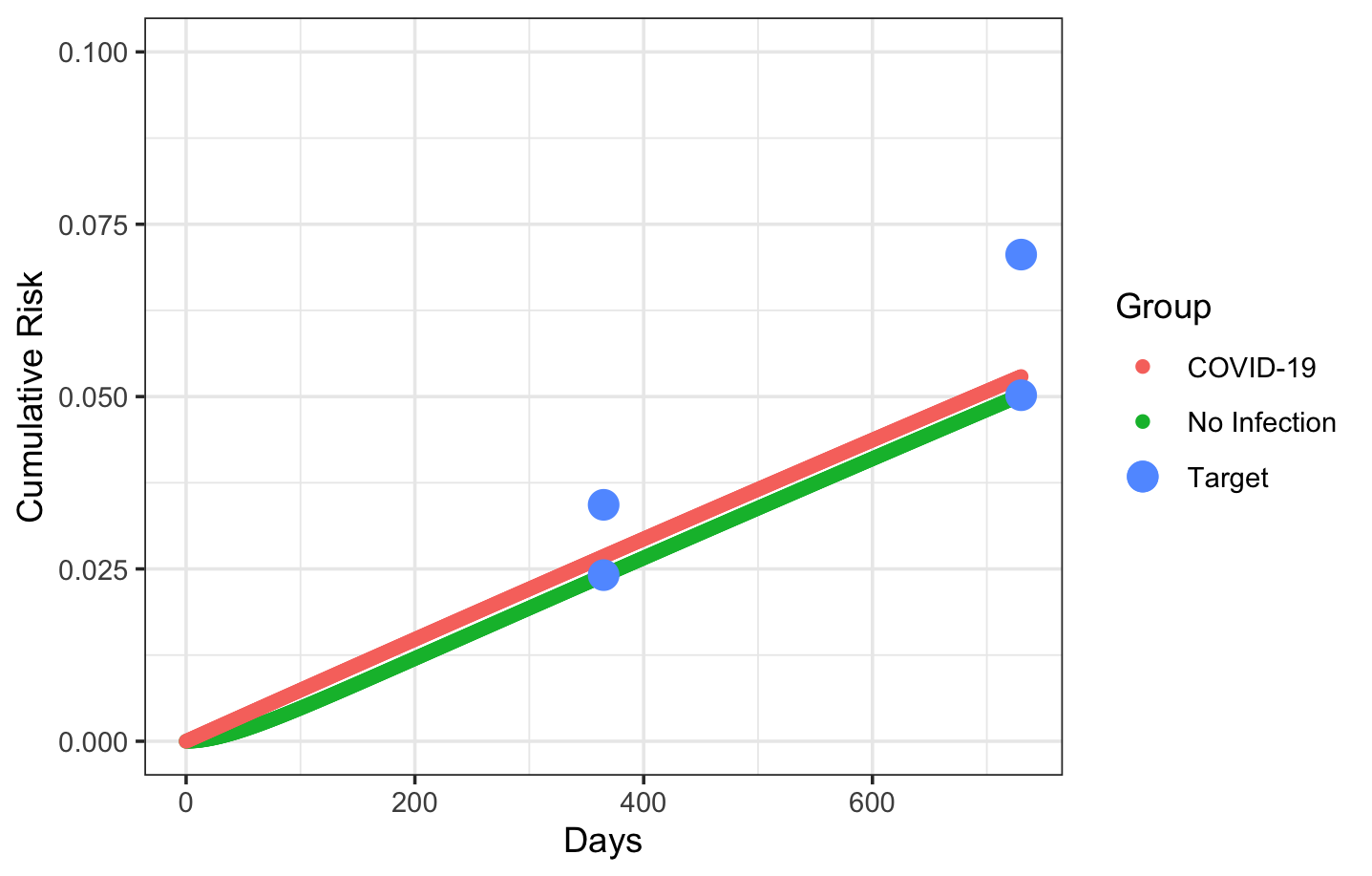

*Second bound: Smallest effect on incident HTN with largest observed HTN diagnostic confound*

In a similar exercise, we can fix ${HRR}_{2}$ at the estimated value of the effect of COVID-19 on changes in diagnostic testing rate, and then we solve for the smallest value of ${HRR}_{1}$ that makes the model consistent with the differences in the cumulative risk of diagnosed HTN at 2 years. If the smallest value of is substantially greater than 1, then we have an estimate of a non-null causal effect of COVID-19 on incident hypertension after removing the confounding effect of changes in diagnostic intensity.

If we fix the underlying rate of incident hypertension for before COVID-19 infection ($r_{1}=0.000075$ per day) and also fix the rate of HTN diagnostic testing before COVID-19 infection ($r_{2}=0.02633$ per day or 0.79 per month) and the increase in HTN diagnostic testing post COVID-19 to be ${HRR}_{2}=$1.26, we can then solve for the smallest increase in incident HTN due to COVID-19 (${HRR}_{1}$), which is 1.399, substantially greater than a null causal effect. Under these assumptions, the model and target data are highly consistent.

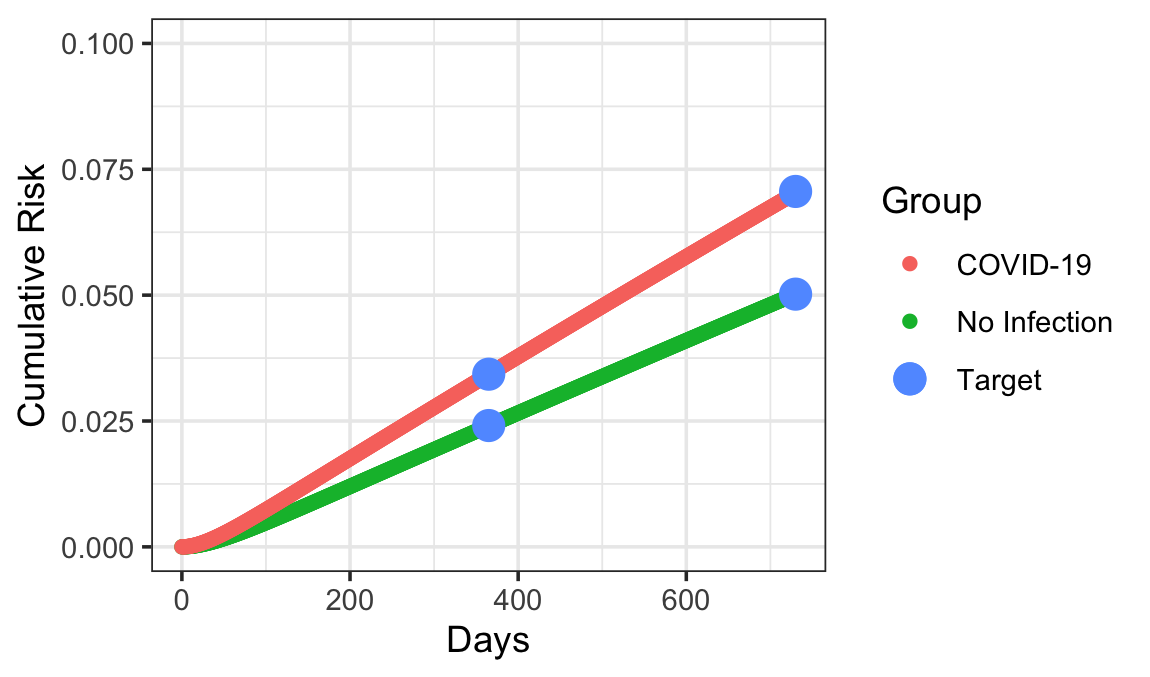

The figure below shows likely values of r1 and HRR1 when the increase in HTN diagnostic testing due to COVID-19 is between 1.26 and 1.28. In it, HRR1 is very unlikely to be below 1.25 let alone 1.0.

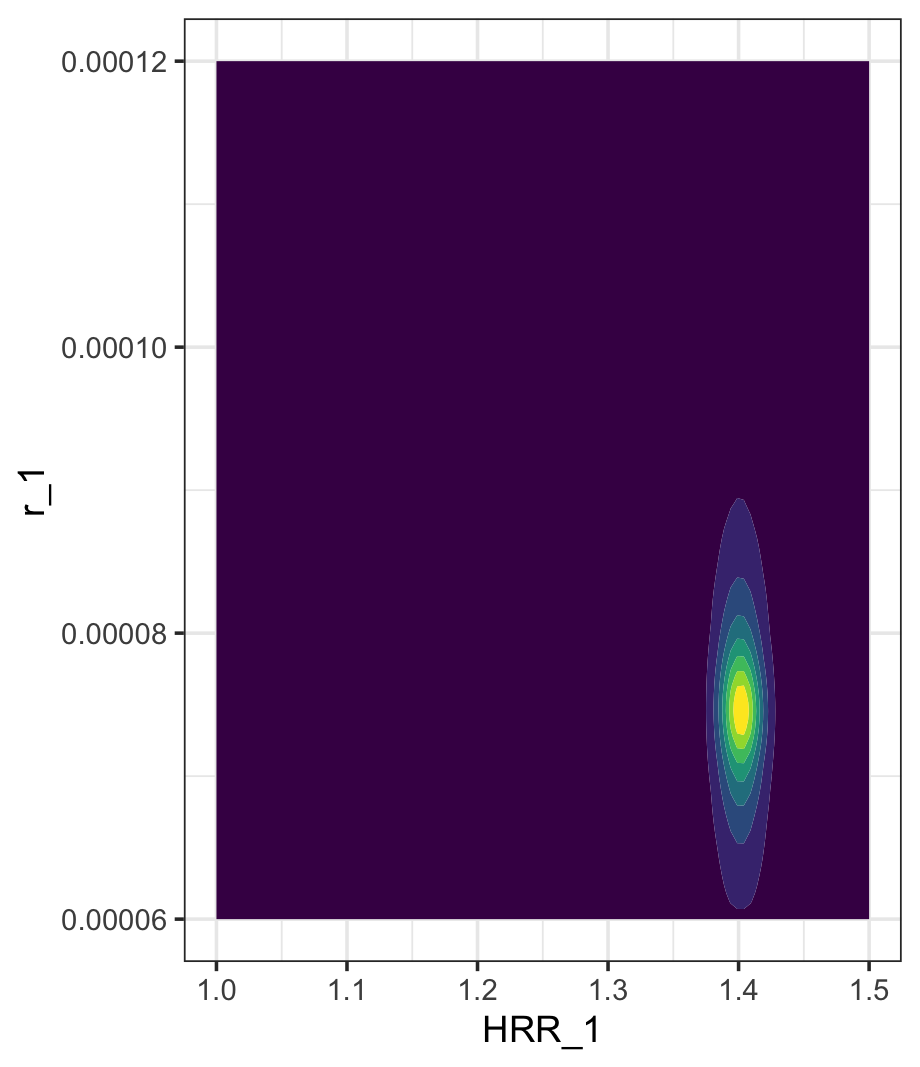

*Overall Conclusion*

Based on these two exercises, we conclude that even after accounting for the confounding effect of increased HTN diagnostic testing rates due to COVID-19, COVID-19 causally increases HTN incidence rates by a hazard rate ratio of 1.399.

**Appendix Table 1. Results for Estimates and Tests of Significance for Interacted Effects (Estimates are Log HRRs: the Sums of the COVID-19 Main Effect and the Relevant Interaction Effect)**

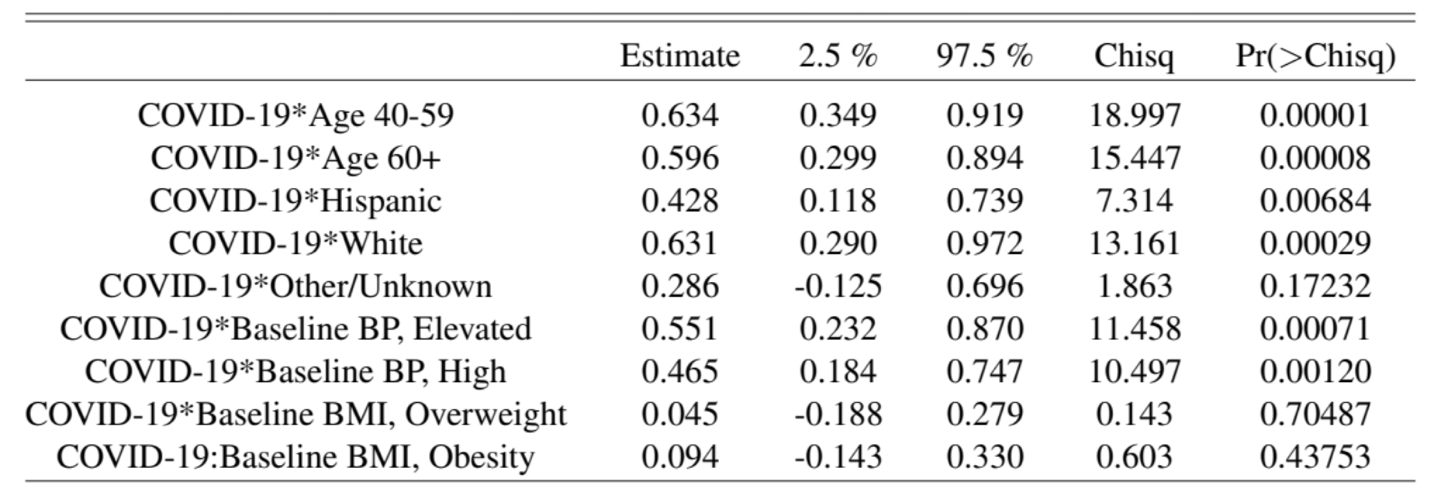

**Appendix Table 2. Predicted Cumulative Risks of Incident Hypertension at 1 Year and 2 Years after Positive COVID-19 Test**

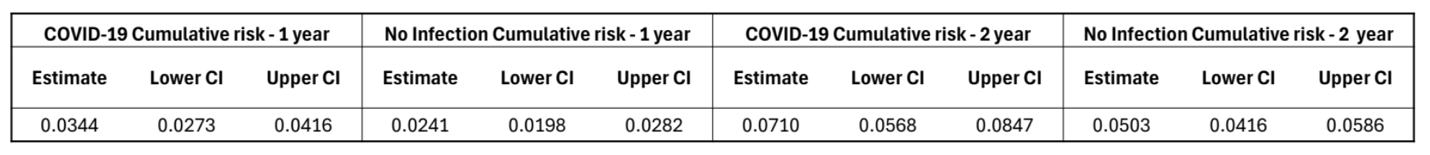

**Appendix Table 3. Predicted Cumulative Risks of Incident Hypertension at 1 Year and 2 Years after Positive COVID-19 Test by Baseline Blood Pressure and BMI Categories**

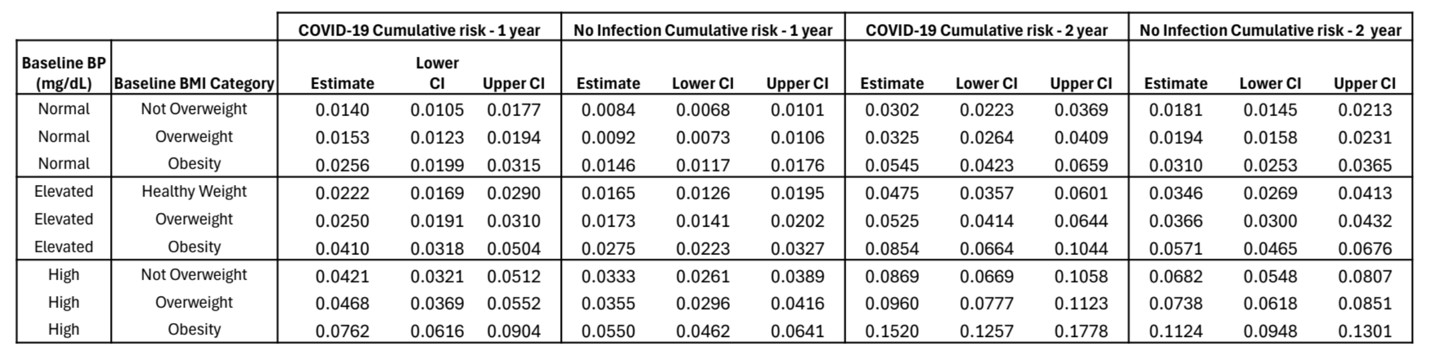

**Appendix Table 4. Predicted Cumulative Risks of Incident Hypertension at 1 Year and 2 Years after Positive COVID-19 Test by Baseline Blood Pressure and Age Categories**

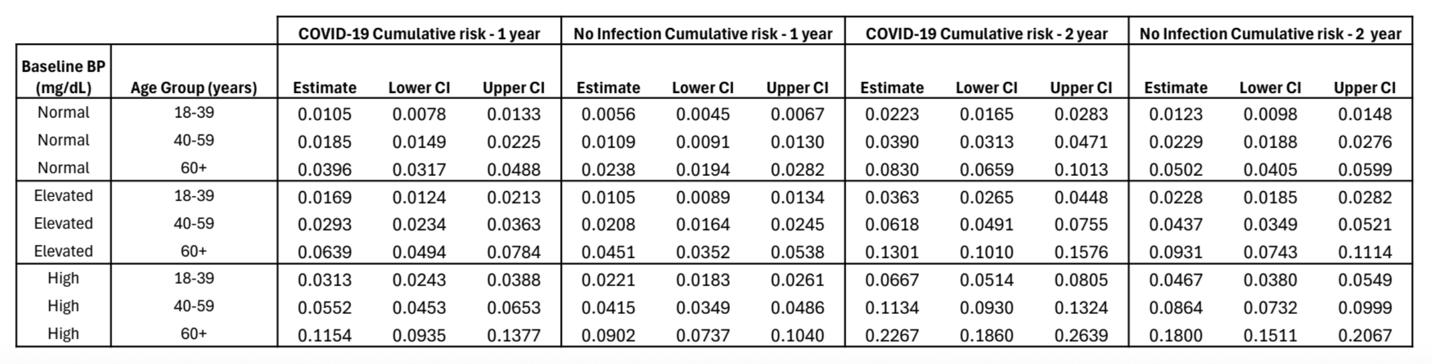

**Appendix Table 5. Baseline Characteristics of Study Population: COVID-19 Test Positive Date Starts Post-Exposure Observation Time**

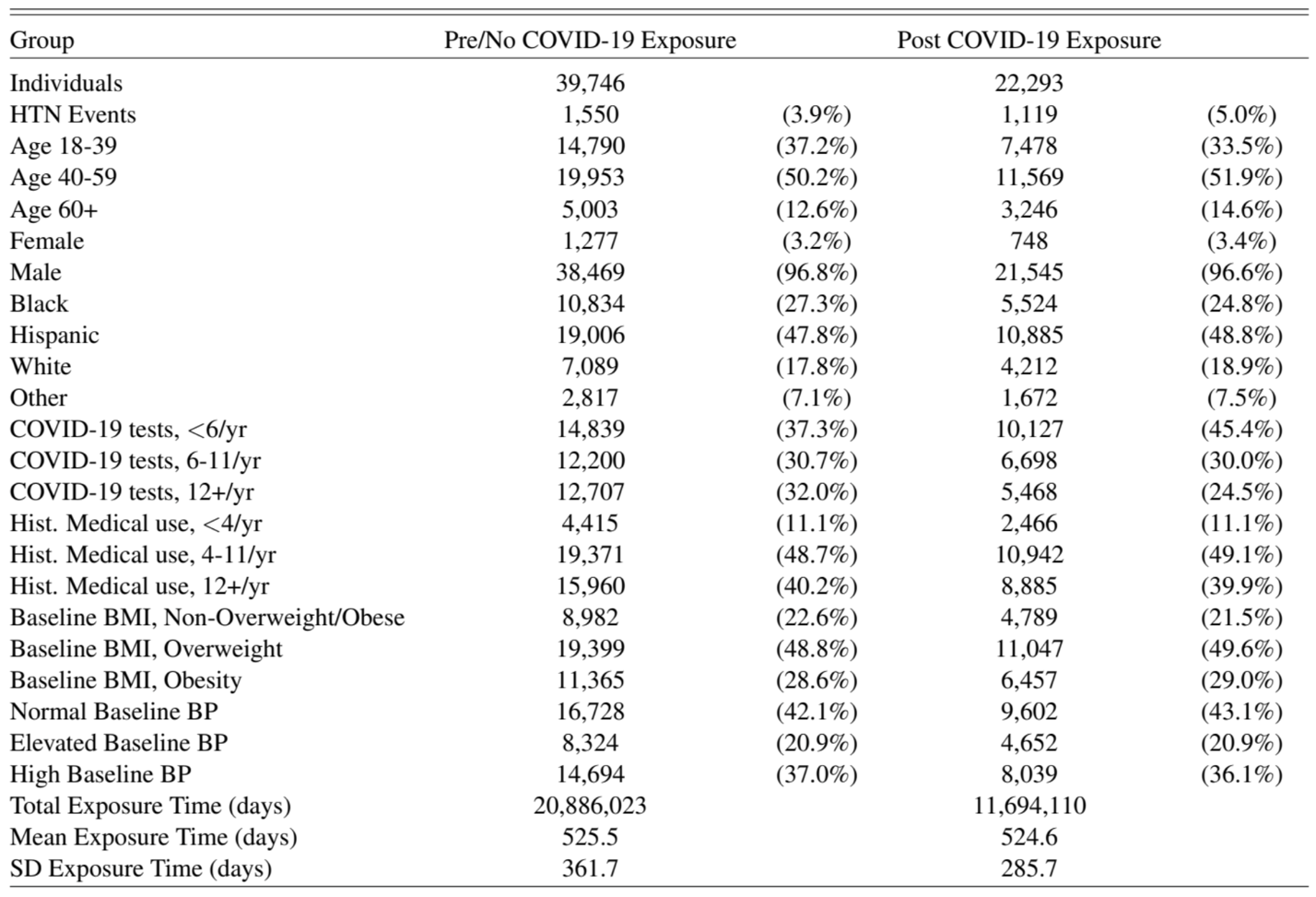

**Appendix Table 6. Baseline Characteristics of Study Population: 61 Days after COVID-19 Test Positive Date Starts Post-Exposure Observation Time**

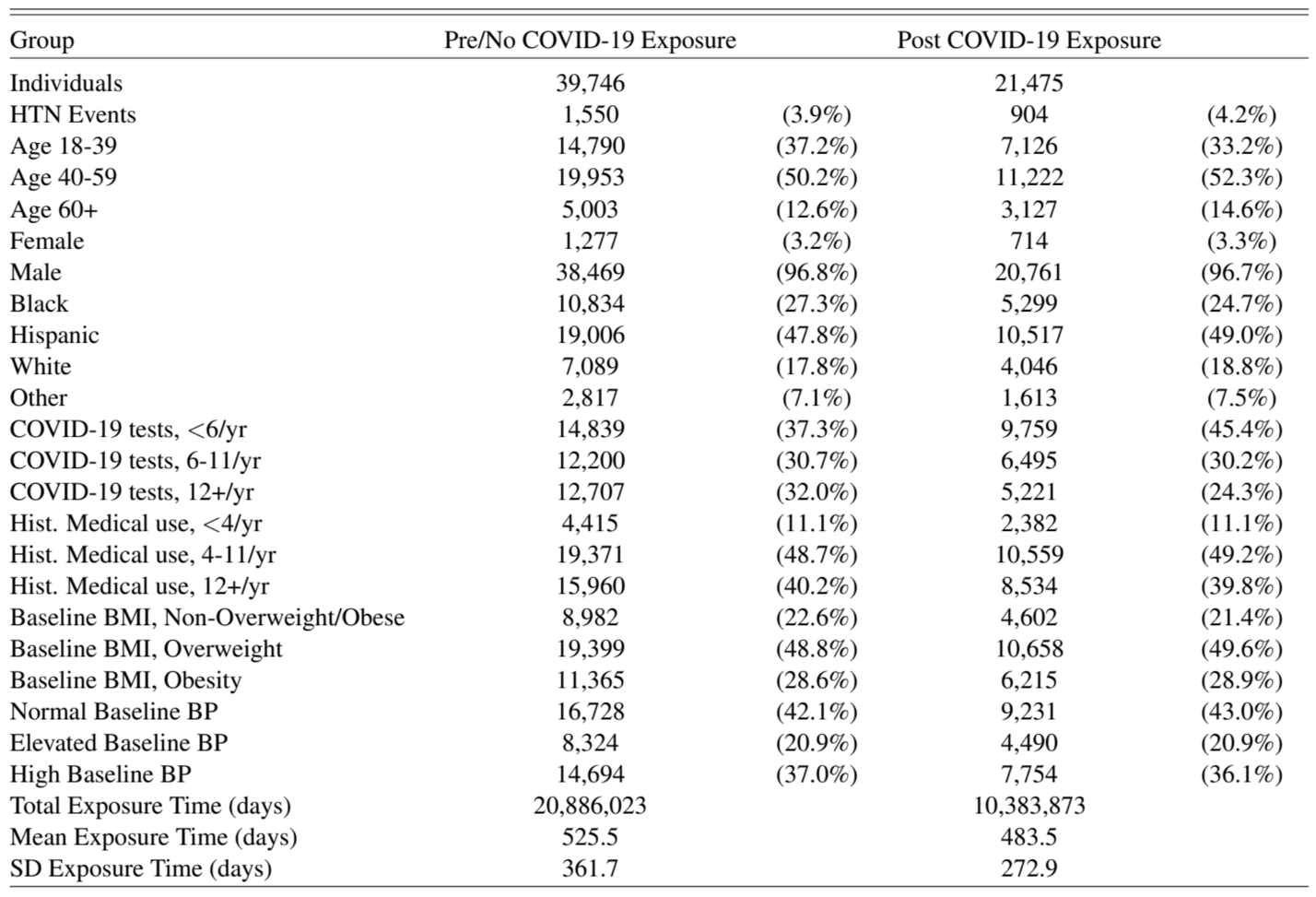

**Appendix Table 7. Baseline Characteristics of Study Population: 91 Days after COVID-19 Test Positive Date Starts Post-Exposure Observation Time**

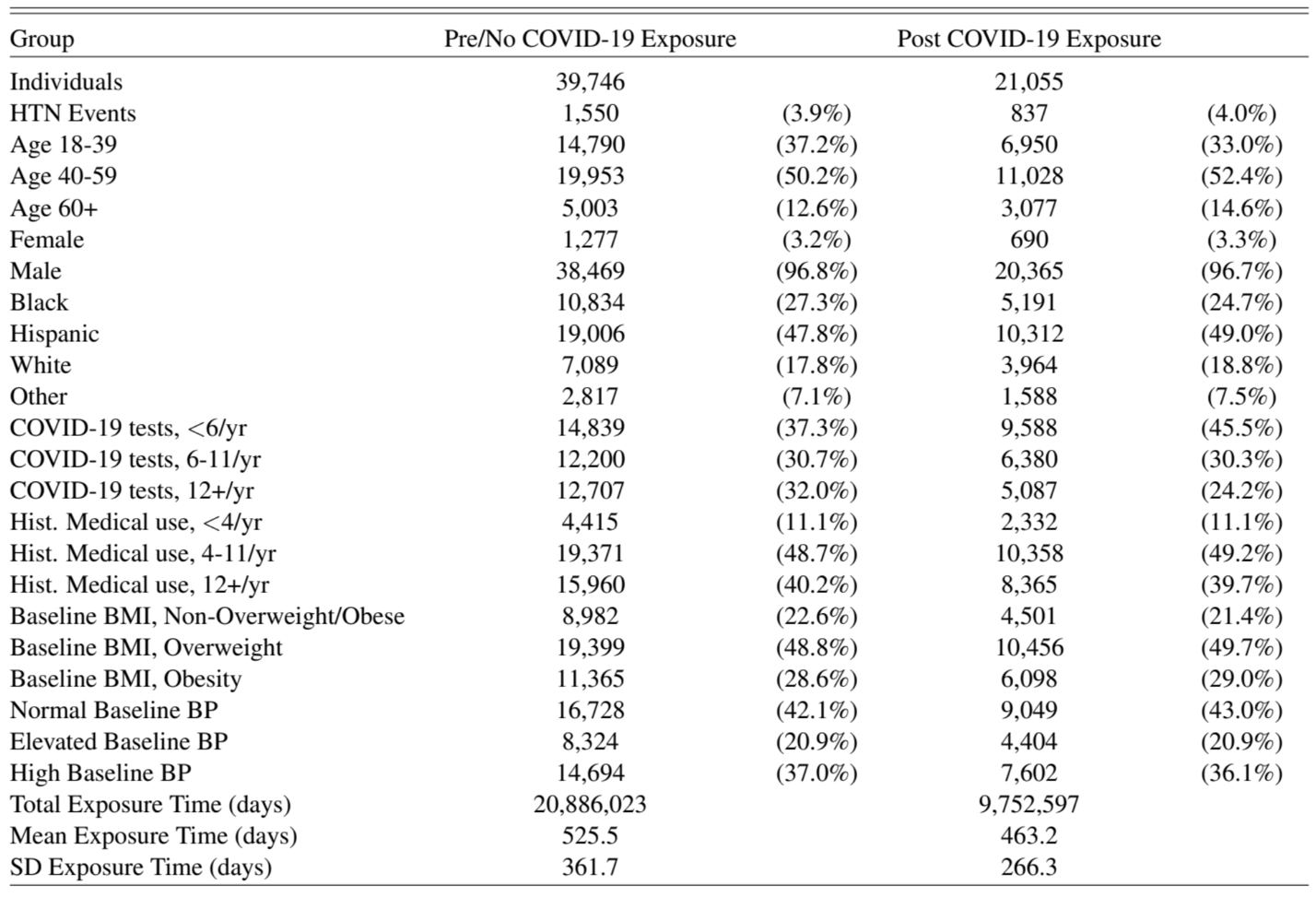

**Appendix Table 8. Multivariate Regression Results for 0-day, 61-day, and 91-day Windows after a Positive COVID-19 Test Starting the Post-exposure Observation Period**

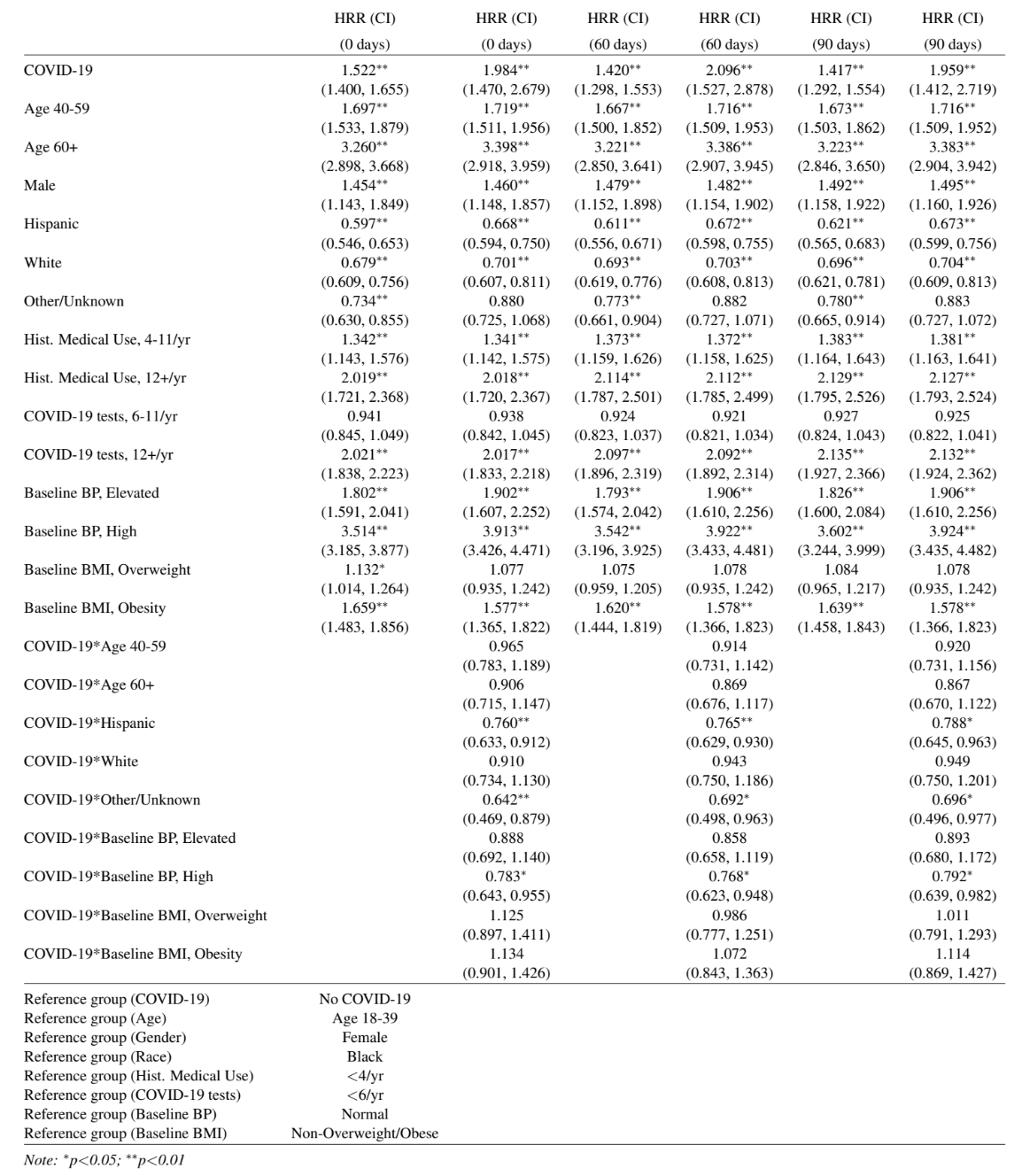
